# Dengue virus infected patients can generate false positives in the ‘gold standard’ *Leptospira* spp. microscopic agglutination test (MAT), immunofluorescence assays (IFAs), and immunoblot assays due to cross-reactive IgG and/or IgM antibodies against their outer surface membrane proteins

**DOI:** 10.1101/2023.03.13.23286524

**Authors:** Claudia M. Romero-Vivas, Margarett Cuello-Pérez, Andrew K. Falconar

## Abstract

There are overlapping world distributions of the mosquito-borne dengue viruses (DENVs) and water-borne bacterial disease leptospirosis which cause large numbers of human infections and fatalities. As such, early differential diagnosis is required for appropriate early leptospirosis antibiotic therapy or DENV patient supportive care, but co-infections have also been reported. From 200 paired (S1 and S2) serum samples collected from suspected DENV infected patients, 70 (35%) were confirmed as ‘on-going’ infections by demonstrating ≥ 4-fold S1 to S2 sample increased anti-DENV IgG and/or IgM ELISA titers. Of those, 8.57% (6/70) also displayed ≥ 4-fold increased S1 to S2 sample titers in the ‘gold standard’ *Leptospira* spp. microscopic agglutination test (MAT) and paraformaldehyde (cell-membrane-impermeable fixative) treated leptospires in immunofluorescence assays (IFAs) due to cross-reactions with 68-72 and 38-42 KDa outer surface membrane antigens present on all *Leptospira* spp. serovars tested. While DENV-1, -2 or -3 serotypes were isolated from their S1 sera: a) *Leptospira* spp. could not be isolated from them, b) their S1 sera were all PCR-negative using a *Leptospira* spp.-specific gene target, and c) their S2 sera were all negative using a commercial anti-*Leptospira* spp. IgM ELISA. As such, we believe this is the first report of DENVs causing false positive reactions in the ‘gold standard’ *Leptospira* spp. MAT, IFAs, and immunoblot assays and which needs further assessments using patients’ samples which were possibly falsely reported as ‘DENV-*Leptospira* spp. co-infections’ in other studies, and to identify these 68-72 and 38-42 KDa *Leptospira* spp. outer surface membrane antigens by proteomic analyses.

## Introduction

Both dengue fever and its more severe forms (severe dengue or dengue hemorrhagic fever/dengue shock syndrome), caused by four different serotypes of flavivirus (DENV-1, DENV-2, DENV-3 and DENV-4) and leptospirosis, caused by different *Leptospira* spp. bacterial serovars, are very important causes of human disease in most tropical and subtropical regions of the world. While the DENVs which do not have important non-human reservoirs are mainly transmitted by the domestic anthropophilic *Aedes aegypti* mosquito species between humans **(1,2)**, a range of pathogenic *Leptospira* spp. serovars are transmitted to humans to mainly through infected urine of different wild and peridomestic (e.g. rats, mice and foxes/stray dogs) and domestic (e.g. dogs, horses, pigs and cattle) animal species **(3)**.

The dengue viruses (DENVs) are now estimated to cause 390-400 million human infections and 21,000-36,000 thousand deaths annually [https://www.cdc.gov/dengue/training/cme/ccm/page51440.html#:~:text=Each%20year%2C%20an%20estimated%20400,deaths%20are%20attributed%20to%20dengue; https://www.who.int/publications/i/item/9789241547871; https://www.worldmosquitoprogram.org/en/learn/mosquito-borne-diseases/dengue], while leptospirosis is now considered to be the most important zoonotic disease of humans with an estimated 1.03 million annual infections resulting in 58,900 deaths **(4)**. As such, leptospirosis is also a highly neglected disease despite its world importance.

Death rates can be dramatically reduced for ‘severe dengue’ (also known as ‘dengue hemorrhagic fever/dengue shock syndrome’: DHF/DSS) to less than 1% through appropriate supportive patient management and which has been achieved in many countries in the world [https://www.who.int/publications/i/item/9789241547871; https://apps.who.int/mediacentre/factsheets/fs117/en/index.html].

Dengue fever (DF) and leptospirosis cases, however, often display similar clinical symptoms and their numbers are often co-elevated throughout these endemic tropical and subtropical regions during times of high rainfall due to i) increased breeding of the *Aedes aegypti* vector species **(1,2)**, and ii) increased contact and infection with leptospires through cuts and mucosa when exposed to contaminated urine present in flooded water **(3)**. As such, differential diagnoses are required to design prompt therapies for them, such as early appropriate antibiotic administration for leptospirosis cases and appropriate supportive care for DENV infected patients.

The ‘gold standard’ serological assay for DENV infections is the plaque reduction neutralization test (PRNT), but is labor intensive and requires specialized equipment and adequately trained staff **(1,2)**. As alternatives: a) paired (S1 and S2) serum/plasma samples obtained 2-14 days apart can be assessed for greater than four-fold increased immunoglobulin M and G (IgM and IgG) titers in ELISAs or hemagglutination inhibition (HAI) assays **(1,2),** b) DENV cDNA can be identified in patients’ acute phase serum samples using DENV (serotype) specific reverse-transcription polymerase chain-reactions (RT-PCRs), or c) DENVs can be isolated and identified be culture and subsequently confirmed using such DENV serotype-specific RT-PCRs or monoclonal antibody (MAb) reactions **(1,2)**. Importantly, while: i) primary DENV cases show a more rapid rise in specific IgM titers with 50% being positive on day 3-5 after the onset of symptoms, ii) secondary DENV cases (due to sequential infections with heterologous DENV serotypes) display a higher and more rapid increase in their specific IgG titers than their IgM titers and many (up to 35%) patients may not even raise detectable IgM titers **(5,6)**.

For the diagnosis of leptospirosis, the ‘gold standard’ is the microscopic agglutination test (MAT) using live or inactivated *Leptospira* spp. cultures of different serovars and the results are determined using dark-field microscopy **(3)**. While in most cases, positive antibody reactions are not detected within one week of the onset of symptoms they may be considered to be positive when serum MAT titers of > 1/100, 1/200, 1/400 or 1/800 are obtained, depending upon the particular laboratory and its location, or when there is a 4-fold or greater increase in their MAT titers between the patients’ first (S1) and second (S2) serum samples obtained 2-14 day apart **(3)** as is also used for DENV diagnosis **(1,2)**. However, many laboratories do not contain the equipment to routinely perform MATs and therefore opt to perform commercially available anti-*Leptospira* spp. specific IgM ELISAs (or rapid lateral flow assays) or specific PCRs on suspected patients’ samples **(3)**.

There have now been many published reports of the need for laboratory tests to differentiate patients with DENV or *Leptospira* spp. infections or cases of dual DENV-*Leptospira* spp. infections. In these studies, performed in many different countries, the DENV diagnoses were determined using: a) different anti-DENV IgM detection (ELISA or rapid lateral flow) assays **(7–14),** anti-DENV IgG detection (ELISA or rapid lateral flow) assays **(8,10,11),** DENV nonstructural-1 (NS1) glycoprotein detection (ELISA or rapid lateral flow) assays **(11)**, RT-PCRs **(8-11,13),** or DENV cell-culture isolation followed by DENV serotype detections **(9)**, while *Leptospira* spp. diagnoses were determined using: b) anti-*Leptospira* spp. IgM (ELISA or rapid lateral flow) assays **(8,11–14),** anti-*Leptospira* spp. IgG detection (ELISA or lateral flow) assays **(11)**, anti-*Leptospira* spp. IgM and IgG IFAs **(7)**, PCRs **(8,10)**, latex agglutination assays **(13)**, or *Leptospira* spp. MATs **(8-10,13-16)**.

We have shown that all four DENV serotypes co-circulate in a major tropical seaport city in the Americas **(5,6)** in addition to the highly pathogenic *Leptospira interrogans* Icterohaemorrhagiae/Copenhageni serovar isolated from a rat (*Rattus rattus*), the *Leptospira interrogans* Pomona serovar isolated from a pig, and the *Leptospia noguchii* Nicaragua and Orleans-like serovar isolates from a dog and water sample, in addition to others serologically identified in humans, dogs, mice, and rats (*Rattus rattus* and *Rattus novegicus*) **(17,18)**.

While very few laboratories routinely collect paired human patients serum samples for DENV diagnosis, we collected 200 paired acute phase S1 and convalescent phase S2 serum samples 2 to 14 days apart from suspected DENV infected patients and perform IgM- and IgG- capture ELISAs to obtain very accurate reciprocal log_10_ 50% end-point ELISA titers (1/log_10_t_50_) through serial dilutions to confirm classical or non-classical primary or secondary DENV infections, together with DENV isolation and serotype determinations **(5,6)**. In this study, we further test the paired serum samples from 70 cases which were confirmed to have on- going DENV infections for their possible co-infection with *Leptospira* spp. using the ‘gold standard’ *Leptospira* spp. MAT using a range of different serovars as well as a specific PCR, IFAs and a commercial anti-*Leptospira* spp. IgM ELISA to identify DENV/*Leptospira* spp. co-infections or any cross-reactions between these DENV infected patients’ IgG and/or IgM antibodies with *Leptospira* spp. antigens which could result in false positive reactions.

## Materials and Methods

### Patients’ samples and data

The collection paired serum samples and clinical data from 200 patients with suspected dengue virus (DENV) infections in the 1.2 million person populated Caribbean Coastal city Barranquilla, Colombia where all four DENV serotypes are endemic, was permitted by the Universidad del Norte Ethics Committee according to National and International guidelines **(5,6)**. Written consent was obtained from all patients or their parents in the case of children before any samples or clinical details were collected from them. These patients’ clinical data and virological, serological, hematological and biochemical results using paired (S1 and S2) serum samples, collected 2-14 days apart according to the WHO guidelines, as was reported **(5,6)**. In this study, these paired (S1 and S2) patients’ serum samples were again assessed for their average IgM and IgG anti-DENV reciprocal log_10_ 50% end-point (1/log_10_t_50_) titers by performing full titrations in our ‘in house’ IgM- and IgG- capture ELISAs to avoid data duplication.

### Growth of dengue viruses

The isolation and growth and serotype determinations of dengue viruses (DEVs) have been described previously **(5,6)**. For growth of each DENV serotype for the IgM and IgG capture (MAC- and GAC-) ELISAs DENV-1 (Nauru Island strain), DENV-2 (Barranquilla S#42 strain) DENV-3 (PR1340 strain) and DENV-4 (Dominica strain) were used to infect 70% confluent C6/36 *Aedes albopictus* cells in 80 cm^2^ flasks containing Leibovitz L-15 medium with 10% v/v tryptose phosphate broth, 10% v/v fetal bovine serum, L-glutamine, sodium pyruvate, and penicillin/streptomycin antibiotics (complete L-15 media: CL-15M). After incubation at 28°C for 4 days, the supernatants were harvested and replaced by fresh CL-15M for incubation for an additional 4 days (day 8). These supernatants were then adjusted to pH 7.2 by an orange-red phenol red indicator color with several drops of sterile 1 M Trisma base/HCl pH 7.2, clarified by centrifugation at 200 x g for 10 min and aliquots were stored at −80°C until required.

For DENV isolations from patients’ sera, 200 μl of their first (S1) serum samples were made to 5 mls with CL- 15M, 0.1 μm syringe-filtered into 25 cm^2^ flasks of 90% confluent C6/36 cells, and incubated for 1 hr at 28°C. These supernatants were then collected and replaced with 5 mls of fresh CL-15M, the collected supernatants were then centrifuged at 200 x g for 10 min, the cell pellet re-suspended in another 5 mls of fresh CL-15M, and then reintroduced into the appropriate flasks (total volume 10 mls per flask) to return the serum-detached cells. After incubation at 28°C, the supernatants were collected and replaced with fresh CL-15M on day 10, 14, 18 and 22. These supernatants were adjusted to pH 7.2 by the addition of a few drops of 1M Trisma base/HCl pH 7.2 to obtain an orange-red phenol red indicator color, clarified by centrifugation at 200 x g, and the supernatant aliquots stored at −80°C. These cell pellets were then re-suspended in 200 μl of PBS and 10 x 10 μl volumes were added to 5 duplicate wells of a 12 well PTFE-coated immunofluorescent slides (Hawksley, UK), air dried before being fixed with cold (−20°C acetone) for 10 min, and air dried again before storage at −40°C until they were required for the immunofluorescent assays (IFAs) (see below).

### Growth of *Leptospira* spp. serovars

The growth of a wide range of *Leptospira* spp. serovars was described **(17)**. For these studies, 19 pathogenic *Leptospira interrogans* Australis, Autumnalis, Ballum, Bataviae, Canicola, Celledoni, Djasiman, Grippotyphosa, Hebdomadis, Icterohaemorrhagiae, Louisiana, Manhao, Panama, Pomona, Pyrogenes, Sarmin, Sejroe, Shermani and Tarassovi serovars, 1 intermediately pathogenic *Leptospira fainei* Hurstbridge serovar and 1 saprophytic/non-pathogenic *Leptospira biflexa* Patoc serovar were grown in Ellinghausen, McCullough, Johnson, Harris-Difco (EMJH) medium (BD Difco, Sparks, MD, USA) containing 400 mg/liter 5-fluorouracil (Ebewe Pharma, Unterach, Austria) at 30°C.

### *Leptospira* spp. microscopic agglutination test (MAT)

The ‘gold standard’ *Leptospira* spp. MAT was performed according to the WHO guidelines **(3)** and, was described previously **(17)**. Briefly, cultures of the different *Leptospira* spp. were grown in EMJH medium up to a concentration of McFarlane (turbidity) 0.5 (approximately 5 x 10^8^/ ml). Each of the S1 and S2 serum samples from the 70 patients confirmed to have on-going primary or secondary DENV infections **(5,6)** were tested for their reactions in the ‘gold standard’ *Leptospira* spp. MAT. For this study, these patients’ serum samples were diluted to 1/5, 1/25 and 1/100 in PBS and 50μl were added to duplicate 50 μl volumes of the non-pathogenic *Leptospira biflexa* Patoc serovar. After incubation at 37°C for 60 minutes, agglutination was observed by dark field microscopy. The patient’s serum samples which caused ≥ 50% agglutination at each of those dilutions were further titrated against duplicate two-fold (1/100-1/1600) serial dilutions of each of the pathogenic serovars and the intermediately pathogenic *Leptospira fainei* Hurstbridge serovar, and the highest MAT titers against a particular serovar with a cut-off value of ≥ 1/100 were reported. Acute human leptospirosis was identified when their first (S1) and second (S2) serum samples increased from negative to ≥ 1/100 or showed a ≥ 4-fold MAT titer rise.

### Anti-*Leptospira* spp. IgM ELISA

The anti-*Leptospira* spp. IgM antibodies in these patients’ second (S2) serum samples which were positive in the MATs were also assessed using a commercial diagnostic SD anti-*Leptospira* spp. IgM ELISA, (Standard Diagnostics, Yongin-si, South Korea), following the manufactureŕs instructions. Positive results were obtained with the average absorbance values were > 0.800 determined at 450 nm, as specified by the manufacturer.

### *Leptospira* spp. *LipL32* gene polymerase chain reaction (PCR) test

The use of the *Leptospira* spp. *LipL32* PCR test was described **(19)**. For this assay, genomic DNA was extracted from duplicate 200 μl volumes of human patients’ first (S1) serum samples using Qiamp DNA mini kits (Qiagen, Hilden, Germany). Subsequently, standard PCRs were performed using 0.1 μM of the *LipL32*/270F (CGCTGAAATGGGAGTTCGTATGATT) and *LipL32*/692R (CCAACAGATGCAACGAAAGATCCTTT) primers with 25 μl volumes containing 2-5 μl of the serum-extracted DNA, 0.25 mM dNTPs (Bioline, Taunton, MA, USA), 1.0 IU of Taq DNA polymerase (Thermo Scientific Fermentas, Waltham, MA, USA) and 3.0 mM MgCl_2_ following: a) an initial reaction step at 95°C for 5 min, b) 35 reaction cycles at 94°C for 1 min and 55°C for 1 minute, and c) a final extension step at 72°C for 5 min. The final products were then subjected to 2% wv agarose gel electrophoresis with the Tris/acetate/EDTA running buffer containing 0.5 mg/ml ethidium bromide and a *LipL32* DNA band of 423 bp was observed in the positive samples using an ultraviolet light transilluminator.

### DENV IgM and IgG capture (MAC- and GAC-) ELISAs

The details of full anti-DENV IgM and IgG capture ELISA titrations has been described **(5,6)**. We repeated these full IgM- and IgG- capture ELISA titrations on these 70 patients confirmed to have on-going primary or secondary DENV infections on their S1 and S2 serum samples to obtain new reciprocal log_10_ 50% end-point (1/log_10_t_50_) titers. For this study, each well of high binding Immulon 4 ELISA plates (Dynatech, USA) were loaded with 50 μl volumes of a 20 μg/ml solution containing either affinity purified goat anti-human IgM Fc5μ or goat anti-human IgG Fcγ (109-005-129 or 109-005-098, Jackson ImmunoResearch, USA) diluted in 0.16%/0.29% (w/v) carbonate/bicarbonate buffer (pH 9.6). After incubation overnight at 4°C, these plates were washed 4 times with PBS containing 0.05% v/v Tween 20 (P1379: Sigma, USA) (PBS/T), and kept damp in PBS/T before 120 μl volumes/well of 1% w/v gelatin in PBS (pH 7.3) were rapidly added using a 12 channel micropipette added and they were incubated at 25°C for 2 hours to block them. After washing the plates 4 times with PBS/T and left slightly damp with PBS/T, 50 μl volumes of three-fold serial dilutions of duplicate patients’ first (S1) and second (S2) serum samples, prepared in PBS/T containing 0.25% w/v gelatin (PBS/T/G) in another plate, were rapidly added using a 12 channel micropipette working up their concentration gradients. After incubation of the sealed plates at 25°C for two hours (or 4°C overnight) they were washed up the antibody concentration-slopes six times with PBS/T to avoid antibody carry-over in their wells. While still remaining damp with PBS/T, 50 μl/well volumes of a 1/3 dilution of an equal mixture of each dengue virus (DENV) serotype from the infected supernatants collected from day 4 or 8 cultures in C6/36 cells were added for the IgM or IgG antibodies to capture the DENVs. After washing the plates 6 times with PBS/T, a 50 μl/well volume of a 1 μg/ml of the affinity purified mouse anti-flavivirus E glycoprotein monoclonal antibody (MAb) 2C5.1 **(20)** in PBS/T/G was allowed to react with the captured DENVs. After incubation at 25°C for 2 hours, the plates were washed six times with PBS/T and 50 μl/well volumes of a 1/2000 dilution of peroxidase-labelled affinity purified goat anti-mouse IgG (H&L: heavy and light chain reactive) (115-035-146: Jackson ImmunoResearch, USA) in PBS/T/G. After incubation at 25°C for two hours, the plates were washed six times before 50 μl/well volumes of a substrate containing 0.012% *o*-phenylene diamine 2HCl (P1526: Sigma, USA) in 0.025M citric acid/0.05 M Na_2_HPO_4_ buffer (pH 5.0) with 0.006% v/v hydrogen peroxide (H1009: Sigma, USA) (opd substrate) and incubated in the dark for 10 minutes before: a) being treated with a 25 μl/well volume of 2M H_2_SO_4_, b) their absorbance values being determined at dual 490 nm with reference 630 nm wavelengths using a 96-well plate reader (MRX: Dynex, USA), and c) their reciprocal log_10_ 50 % end-point titers (1/log_10_t_50_) were determined from their average 50% maximum 490 nm absorbance values (Abs Max/2). Primary DENV infections (P) were serologically confirmed by observing a greater than 4-fold increase in their IgM titers and which were higher than their IgG titers between the patient’s S1 and S2 serum samples while secondary DENV infections (S) were determined by observing a greater than 4-fold higher IgG titers, and which were higher than their IgM titers between the patient’s S1 and S2 serum samples. Any ‘non-classical’ (*S, *P, *LS or *PS) anti- DENV antibody reactions were also identified as described previously **(5,6).**

### Immunofluorescence assays (IFAs) of DENV serotypes isolated from patients or patients’ IgM and IgG antibodies against *Leptospira* spp. outer surface membrane proteins

The immunofluorescence assays (IFAs) used to identify the DENV serotypes isolated from patients’ S1 serum samples after infection of the C6/36 *Aedes albopictus* cell line have been described **(5,6)**. Briefly, the acetone- fixed C6/36 cell pellet-coated slides prepared using the cells liberated in the CL-15M cultures (see **Growth of dengue viruses**: above) were thawed to 25°C, wetted with PBS before 20 μl volumes of 1/200-1/400 dilutions of immunoaffinity-purified mouse monoclonal antibodies (MAbs) specific for the DENV-1 (MAb 15F3), DENV-2 (MAb 3H5), DENV-3 (MAb 5D4) or DENV-4 (MAb 1H10), or all flavivirus (MAb 2C5.1 or 4G2) main envelope (E) glycoproteins diluted in PBS containing 2% w/v milk powder (Marvel, Cadbury’s, UK) (PBS/M) were reacted with each pair of these wells at 28°C for 2 hours. After washing 3 times with PBS, 20 μl volumes of a 1/1000 dilution of FITC-labelled goat anti-mouse IgG (H&L) (115-095-062: Jackson ImmunoResearch, USA) diluted in PBS/M were reacted with each well of these slides. After washing 4 times with PBS and very brief inserted into distilled water, the slides wells were lightly blotted before they were mounted under coverslips using 90% glycerine/PBS pH 8.5-8.8. These slides were then viewed under immunofluorescence microscopy using the correct FITC excitation and extinction filters and the DENV serotypes were determined.

The immunofluorescence assays (IFAs) were prepared using 2% w/v paraformaldehyde-fixed *Leptospira* spp. which inhibited patients’ PAbs and the labelled secondary antibodies from entering the cells, as previously reported **(21**: https://www.jove.com/es/t/2805/immuno-fluorescence-assay-of-leptospiral-surface-exposed-proteins). For these assays, cultures of the *Leptospira interrogans* Icterrohaemorrhagiae, Canicola, Pomona and Tarassovi, *Leptospira fainei* Hurstbridge and *Leptospira biflexa* serovars, grown in EMJH medium at 28°C which reached a McFarlane 0.5 (approximately 1 x 10^8^/ml) level, were collected and centrifuged at 2,000 x g for 7 minutes at 25°C. These pellets were then re-suspended in PBS containing 5 mM MgCl_2_ (PBS/Mg^2+^) at pH 7.2 to obtain a final concentration of 1 x 10^8^ leptospires/ml as determined using an improved Neubauer hematocytometer slide. Aliquots were then added to wells in 12-well PTFE-coated immunofluorescent slides (Hawksley, UK) and allowed to adhere for 30 min at 30°C. The supernatants were then carefully removed and the cells fixed with 2% wt/vol paraformaldehyde in PBS/Mg^2+^ pH 7.2 for 40 min at 30°C**. [N.B. This solution was prepared by heating 2% wt/vol paraformaldehyde in PBS at 60°C after being adjusted to pH 9.0 until it dissolved and then adding stock MgCl_2_ solution to obtain a 5 mM final concentration and then slowly (dropwise) adding 1 M HCl to obtain pH 7.2]**. After removal of the fixing solution, these slides were washed 3 times with PBS/Mg^2+^ and then gently blotted before being blocked with EMJH medium for 90 min at 30°C. After briefly washing the slides 4 times with PBS/Mg^2+^, the human S2 (secondary) serum samples from i) Group A (DENV-*Leptospira* spp. co-seropositive) patient A.1 to A.6, ii) Group B (DENV seropositive only: i.e. anti-*Leptospira* spp. negative control) patient B1, B.2 and B.3, and ii) Group C (*Leptospira* spp. seropositive only: i.e. anti-*Leptospira* positive control) patient C.1, C.2 and C.3 **(Table 1)** were diluted at 1/200 in PBS/Mg^2+^ containing 0.25% v/v EMJH medium (PBS/Mg^2+^/E) pH 7.2 and reacted with the slides at 30°C for 60 min. After washing these slides again, a 1/1000 dilution of either FITC-labelled goat anti-human IgM Fc5μ or goat anti-human IgG Fcγ (109-095-043 or 109-095-098: Jackson ImmunoResearch, USA) secondary antibodies diluted in PBS/Mg^2+^/E were added to the appropriate wells and incubated at 30°C for 60 min before washing 4 times with PBS/Mg^2+^ and briefly dipped in distilled water. The slides were then gently blotted and then mounted with PBS/Mg^2+^ containing 90% v/v glycerin pH 9.0 under coverslips and viewed using a fluorescence microscopy with the appropriate FITC excitation and blocking filters and the results were photographed at a 200x magnification.

**Table 1.**
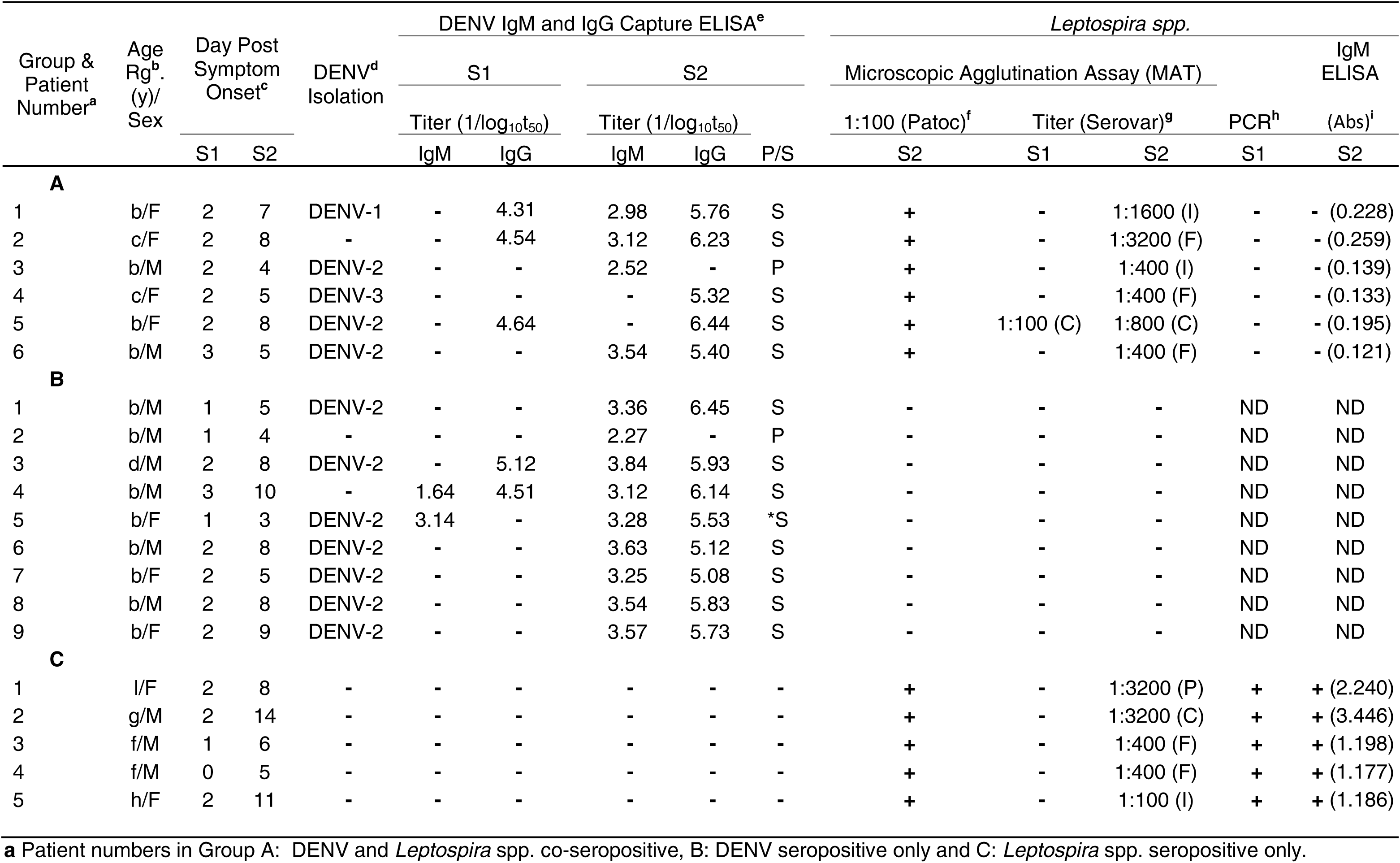
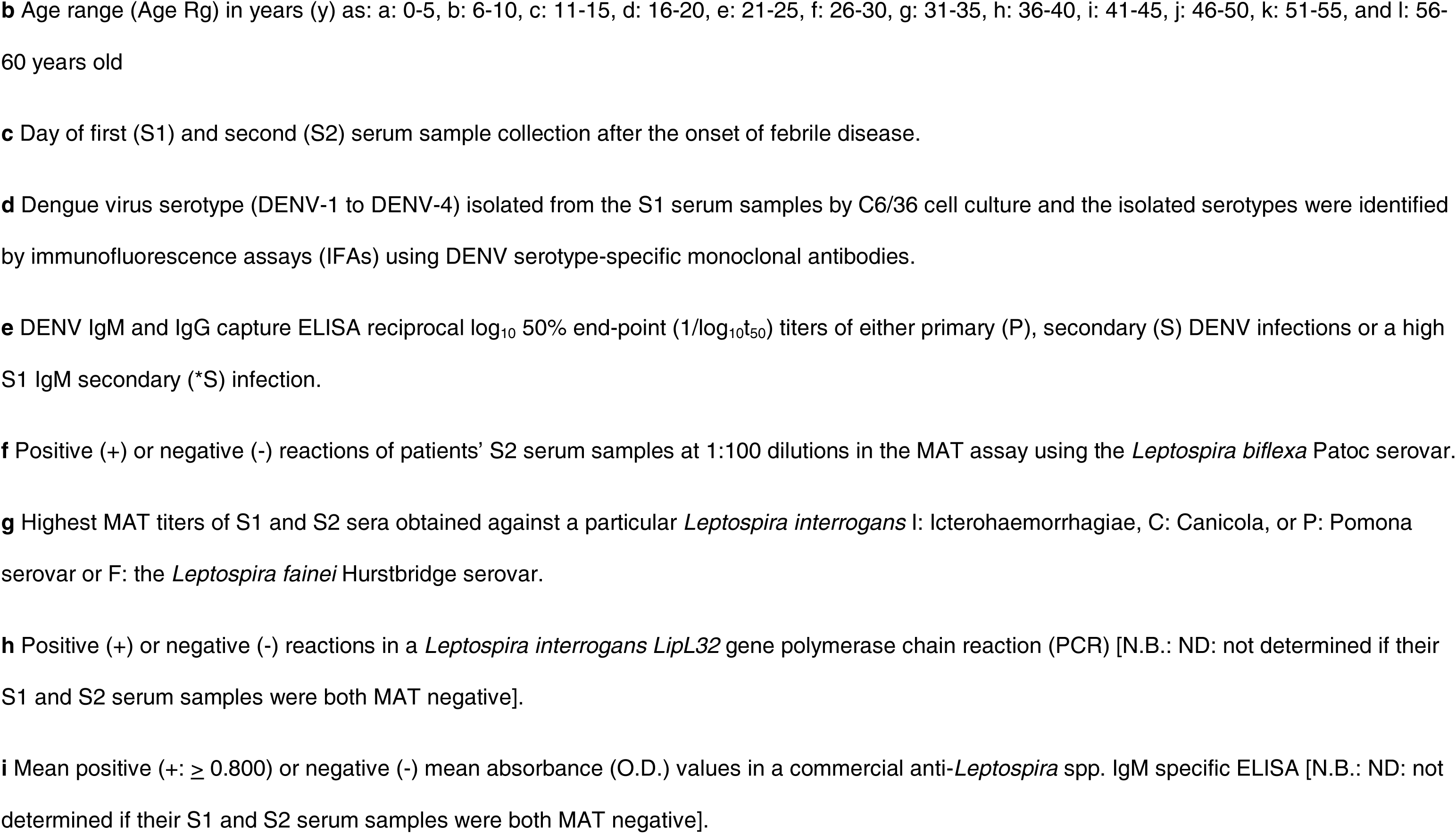
Details of DENV and *Leptospira* spp. co-seropositive patients’ Group A versus patients’ groups which were either only DENV (Group B) or only *Leptospira* spp. (Group C) seropositive.

### Immunoblot (Western blot) assays of patients’ S2 serum samples against *Leptospira* spp. lysates

Alternatively, the *Leptospira interrogans* Icterohaemorrhagiae, Tarassovi, Canicola and Pomona, *Leptospira fainei* Hurstbridge and *Leptospira biflexa* Patoc serovars were concentrated by continuous washing and centrifugation with PBS/Mg^2+^, and then their supernatants were removed and approximately 0.8 x 10^9^ *Leptospira* spp. were lysed in 100 μl volumes of a non-reducing buffer containing sodium dodecyl sulfate (SDS) in buffer at pH 6.8 (33 mM ortho-phosphoric acid/0.62 mM Trisma base and 69.3 mM sodium dodecyl sulfate) at 28°C for 60 min with gentle agitation using a pipette tip to shear the DNA before centrifugation at 2,000 x g to pellet the insoluble components. The clarified supernatants were then collected and aliquots stored at −80°C. These samples were then thawed and 10 μl aliquots were either left non-reduced or reduced by the addition of a 0.5 μl volume of 2-mercaptoethanol (2-ME) (63689, Sigma Aldrich, USA), together with 2 μl/tube of 2.5% w/v bromophenol blue/50% v/v glycerol in H_2_O and heated in a boiling water bath for 3 minutes. After cooling and centrifugation, 10 μl aliquots were added to lanes of a 10-well stacking gel in 9% discontinuous SDS-PAGE gels (Mini Protean II Vertical SDS-PAGE System, BioRad, USA) with wide ranging (8-220 KDa) colored molecular weight markers (C1992: Sigma Aldrich, USA) in the outer lanes. These gels were subjected to constant 200V electrophoresis for approximately 60 min in 0.8% w/v Trisma base (T6066: Sigma Aldrich, USA) and 2.8% w/v glycine (pH 8.7-8.9) electrode buffer. Alternatively, 2-4 well combs were used to prepare preparative nitrocellulose-membrane immunoblot strips (see below), and which were flanked by two 3 mm molecular marker lanes.

These gels were then subjected to immunoblotting (western blotting) using a ‘sandwich’ consisting of a 0.1 μm pore-sized nitrocellulose membrane (Protran BA79: Whatman, USA) flanked by three layers of gel blotting paper (GB005: Whatman, USA) contained in 48 mM Trisma base/39 mM glycine with 40% methanol at 160 mA/gel in a semi-dry electroblot apparatus (Sartoblot II: Sartorius, USA) for 30 min. They were then air-dried before being blocked in a solution of 2-3% milk powder (KLIM, Nestle’s, USA) in PBS containing 0.05% w/v NaN_3_ overnight. After washing the membranes with PBS/T (see above) three times they were either dried as an entire gel blot, or for cutting 1.5-2.0 mm wide preparative strips. After briefly wetting the blots/strips with PBS/T, 1/250 dilutions of human patients’ S2 (secondary) serum samples from: i) Group A (DENV-Leptospira spp. co-seropositive) patient A.1 to A.6, ii) Group B (DENV seropositive only: i.e. anti-*Leptospira* spp. negative control) patient B.1 and Group C (*Leptospira* spp. seropositive only: i.e. anti-*Leptospira* spp. positive control) patient C.1 dissolved in PBS/T containing 2% w/v milk powder (PBS/T/M) (6 ml volumes for entire blots or 1 ml volumes for 1.5-2.0 mm strips) were reacted with these blots at 25°C for 2 hours. The blots/strips were then washed 4 times with PBS/T before a 1/2000 dilution of peroxidase-labelled goat anti-human IgG (H&L: heavy and light chain reactive) (109-035-088: Jackson ImmunoResearch, USA) secondary antibody diluted in PBS/T/M was reacted with them at 25°C for 2 hours before being washed again 4 times with PBS/T and then two times in PBS. They were then incubated with a mixture of 10 mg of 3, 3’ diaminobenzidine (D8001: Sigma Aldrich, USA) pre-dissolved in 5 mls of methanol before adding to 40 mls of PBS containing 30 mg of 4-chloro-1-naphthol (C8890: Sigma Aldrich, USA) (DAB/4C1N substrate). After incubation for 10-30 minutes, these blots/strips were thoroughly washed with distilled water and air dried. They were then photographed in RAW with a 100 mm Canon macro lens attached to a Canon EOS digital camera and then arranged and labelled in PowerPoint.

## Results

From a study of paired (S1 and S2) serum samples from patients with either DENV **(5,6)** or *Leptospira* spp. **(17,18)** infections in our study site, 70/200 patients were again confirmed to have on-going primary or secondary DENV infections by obtaining ≥ 4-fold increase in their reciprocal log_10_ 50% end-point (1/log_10_t_50_) IgG and/or IgM ELISA titers between their S1 and S2 serum samples **(5,6)**, while 16 patients from another study were serologically identified as *Leptospira* spp. seropositive only patients **(17)**. Amongst these 70 confirmed DENV infected patients, 8.6% (6/70) of them (patient A.1 to A.6) were also shown to have on-going *Leptospira* spp. infections by obtaining greater than 4-fold increased S1 to S2 serological titers in the ‘gold standard’ *Leptospira* spp. microscopic agglutination test (MAT) **(Table 1)**. As such, these six patients (patient A.1 to A.6) were included in a Group A for comparison with other ‘control’ patients who were either DENV seropositive only (Group B: n = 9: patient B.1 to B.9) or *Leptospira* spp. seropositive only (Group C: n = 5: patient C.1 to C.5). Three different DENV serotypes (DENV-1, DENV-2 and DENV-3) were isolated by C6/36 cell culture and serotype-specific monoclonal antibody (MAb) determinations from the first (S1) phase serum samples. Of these six Group A (DENV-*Leptospira* spp. co-seropositive) patients, only one of them (patient A.3) had a primary DENV infection as determined by their high IgM- versus lower IgG- capture ELISA titers, they had a 6-30 (b-f) group age range and 2/6 were male patients. In contrast, only DENV-2 was isolated from Group B patients (patients B.1, B.3 and B.5-B.9), but while members of this group had a 6-20 (b-d) group age range, slightly older patients with a 26-60 (f-l) age range were present in Group C. These results were consistent with young children being the most frequent to develop DENV infections in DENV highly endemic areas, while leptospirosis being an occupational disease of adults **(1–3)**, and who were also likely to have previously encountered previous DENV infections.

All Group A and C patients’ secondary (S2) serum samples were positive at 1:100 dilutions against the non-pathogenic *Leptospira biflexa* Patoc serovar in the ‘gold standard’ MAT, while none of the Group B (DENV reactive only: i.e. anti-*Leptospira* spp. negative control) patients’ S2 serum samples were positive against any of these *Leptospira* spp. serovars. The highest MAT titers of the S2 serum samples from Group A and C patients were obtained against the pathogenic *Leptospira interrogans* Icterohaemorrhagiae, Canicola or Pomona serovars, as well as the intermediately pathogenic *Leptospira fainei* Hurstbridge serovar and, very importantly, all of the Group A and C patients’ serum samples showed increased *Leptospira* spp. ‘gold standard’ MAT titers from negative to 1:100 (patient C.5) or 1:400 up to 1:3200 (≥ 4-fold increase).

Amongst the samples from the Group A, B and C patients’ groups: i) only the S1 serum samples from Group C (*Leptospira* spp. seropositive only) patient C.1 to C.5 were positive on the PCR for the *Leptospira* spp.-specific *LipL32* gene target and ii) only their S2 serum samples were positive in the commercial anti-*Leptospira* spp. IgM ELISA. [N.B. Group B patients’ S1 or S2 serum samples were not tested in either the *Leptospira* spp. *LipL32* PCR or commercial anti-*Leptospira* spp. IgM ELISA since all their S1 and S2 serum samples were completely negative in the ‘gold standard’ *Leptospira* spp. MATs].

As such, while the patients from Group A all showed increased S1 to S2 serum sample titers against DENVs in the IgM- and/or IgG- capture ELISAs and *Leptospira* spp. serovars in the ‘gold standard’ MATs, their S1 serum samples were all *Leptospira interrogans LipL32* gene-negative by PCR, and their S2 serum samples were all negative in the commercial anti-*Leptospira* spp. IgM ELISA **(Table 1)**.

### Patients’ IgM and IgG reactions against paraformaldehyde-fixed *Leptospira interrogans* in immunofluorescent assays (IFAs)

Group A (DENV-*Leptospira* spp. co-seropositive) patients’ S2 serum samples were initially tested at one dilution for their reactions against outer membrane antigens of the *Leptospira interrogans* Icterrohaemorhagiae serovar which had been fixed with 2% w/v paraformaldehyde to prevent antibodies from entering the bacterial cells for comparison with the reactions of some Group B (DENV seropositive only) and Group C (*Leptospira* spp. seropositive only) patients’ S2 serum samples **(Figure 1 and 2)**. In this study, the serum samples from Group A patient A.1 and A.2 provided positive IgM and/or IgG reactions against these fixed *Leptospira interrogans* Icterrohaemorrhagiae bacteria in the IFAs **(Figure 1)** as they also did in the MAT, and despite being negative in the commercial anti-*Leptospira* spp. IgM ELISA **(Table 1)**. The S2 serum samples from Group A patient A.3 was however only IgM positive in this IFA **(Figure 1)**, since this patient had a primary DENV infection (i.e. only anti-DENV IgM positive in the DENV IgM- and IgG- capture ELISAs), while also being negative in the *Leptospira* spp. *LipL32* gene PCR and the anti-*Leptospira* spp. IgM ELISA **(Table 1)**. In contrast, the S2 serum sample from Group B (DENV seropositive only: i.e. anti-*Leptospira* spp. negative control) patient B.1 was, as expected, IgM- and IgG-negative against these fixed *Leptospira interrogans* bacteria in these IFAs **(Figure 1)**, as it was in the *Leptospira* spp. MAT, the PCR, and the commercial anti- *Leptospira* spp. IgM ELISA **(Table 1)**. The S2 serum sample from Group C (*Leptospira* spp. seropositive only: i.e. anti-*Leptospira* spp. positive control) patient C.1 was however, as expected, IgM- and IgG- positive in these *Leptospira interrogans* bacterial IFAs **(Figure 1)** as it was in the *Leptospira* spp. MAT, and the commercial anti-*Leptospira* spp. IgM ELISA **(Table 1)**.

**Figure 1.**
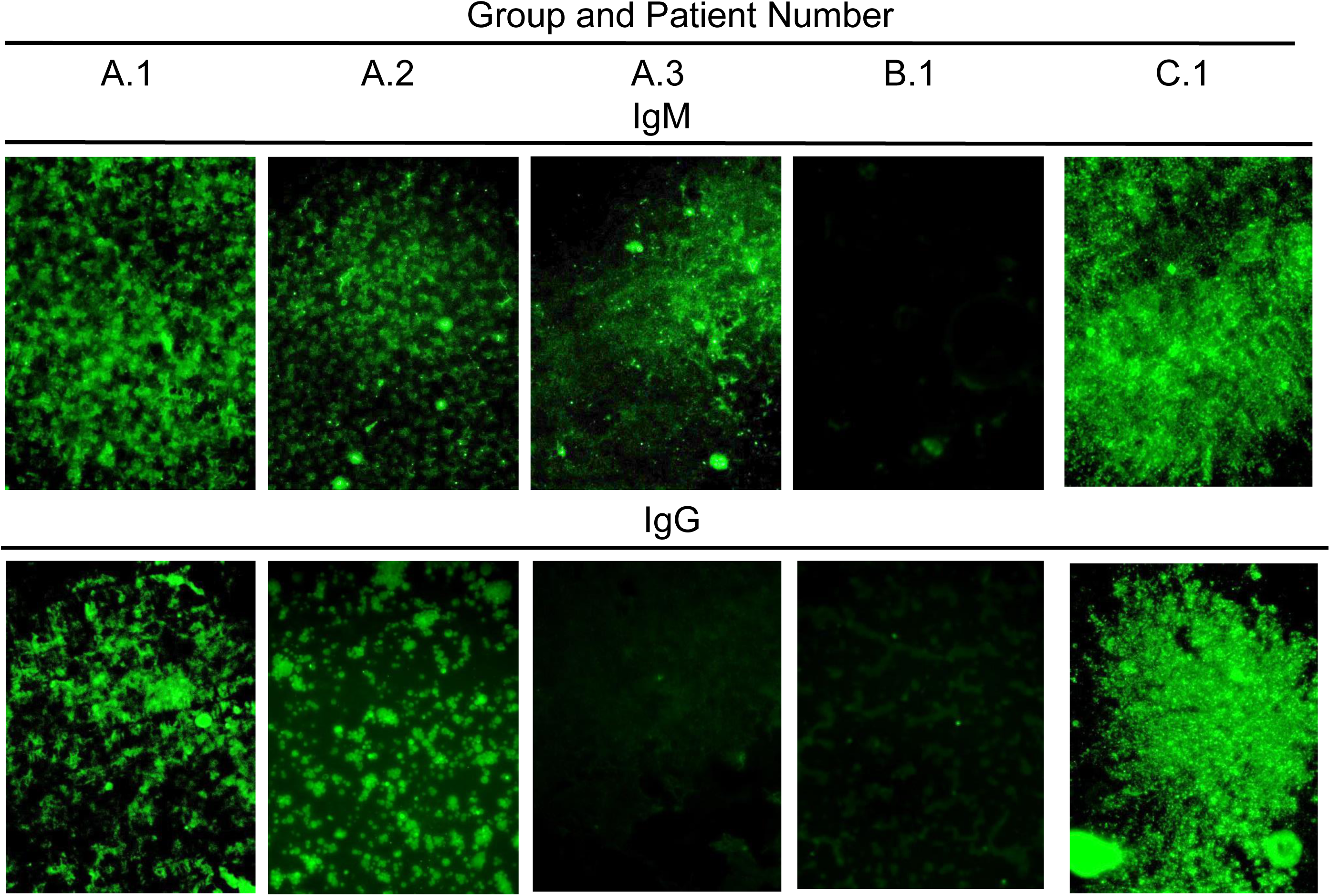
Immunofluorescent assay (IFA) photomicrographs of human patient’s IgM and IgG reactions against paraformaldehyde-fixed *Leptospira* spp. Human S2 serum samples from: i) Group A (DENV-*Leptospira* spp. co-seropositive) patients A.1, A.2 and A.3, ii) Group B (DENV seropositive only: i.e. anti-*Leptospira* spp. negative control) patient B.1 and iii) Group C (*Leptospira* spp. seropositive only: i.e. anti-*Leptospira* spp. positive control) patient C.1 were reacted at a 1/200 dilution with 2% w/v paraformaldehyde-fixed *Leptospira interrogans* Icterrohaemorrhagiae serovar and their reactions were detected using either FITC-labeled anti-human IgM- (IgM) or IgG- (IgG) specific secondary antibodies and all of them were photographed at 200x magnification using immunofluorescent microscopy with the correct excitation and emission filters. [N.B. Since these assays resulted in the formation of large *Leptospira* spp. aggregates, rather than discrete slide-bonded bacteria, the results were clearly either positive (+) or negative (−) at this low 200x magnification and therefore no size bars are shown]

**Figure 2.**
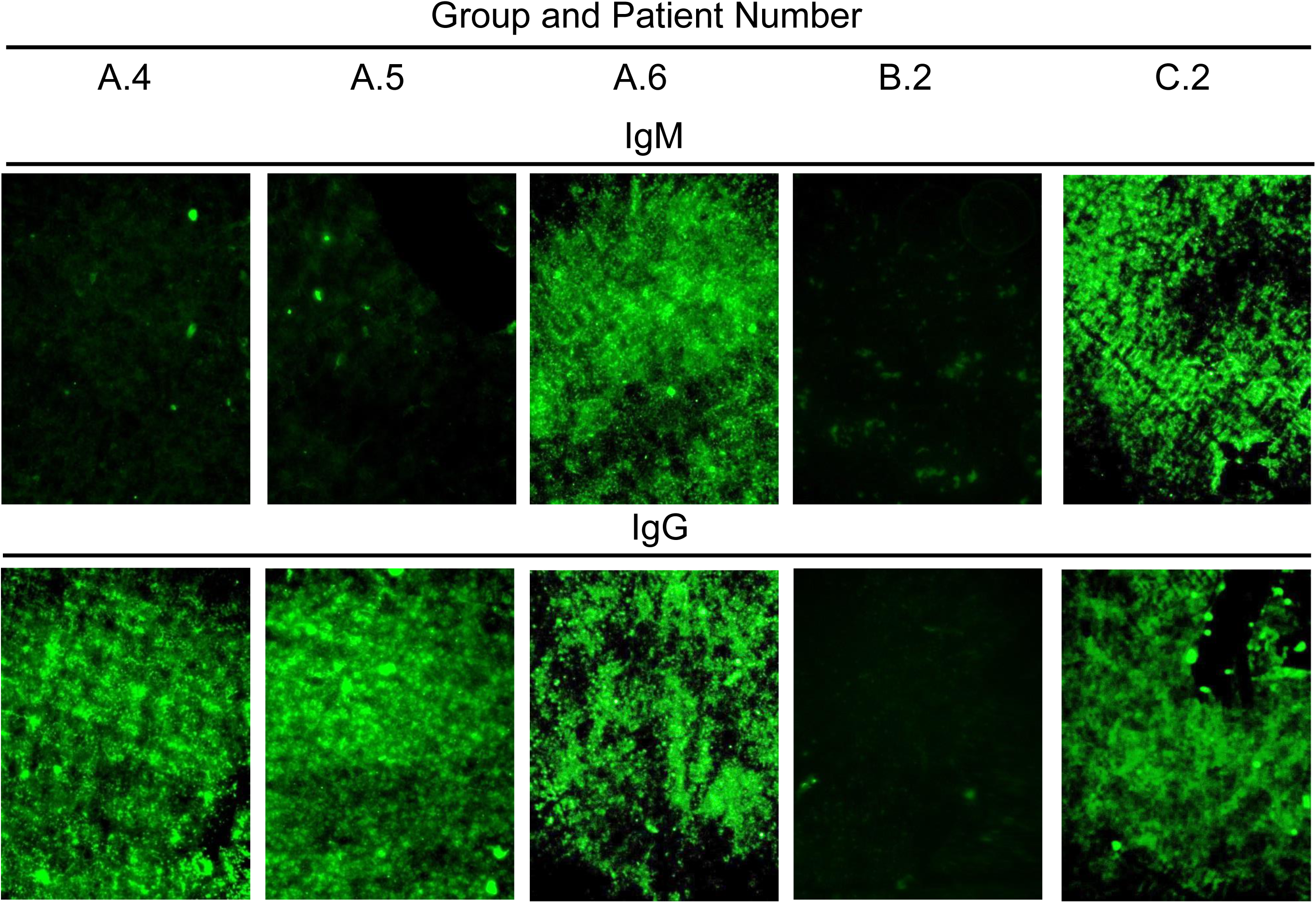
Immunofluorescent test (IFAT) photomicrographs of human patient’s IgM and IgG reactions against paraformaldehyde fixed *Leptospira* spp. Human S2 serum samples from: i) Group A (DENV-*Leptospira* spp. co-seropositive) patients A.4, A.5 and A.6, ii) Group B (DENV seropositive only: i.e. anti-*Leptospira* spp. negative control) patient B.2 and iii) Group C (*Leptospira* spp. seropositive only: i.e. anti-*Leptospira* spp. positive control) patient C.2 were reacted at a 1/200 dilution with 2% w/v paraformaldehyde-fixed *Leptospira interrogans* Icterrohaemorrhagiae and their reactions were detected using either FITC-labeled anti-human IgM- (IgM) or IgG- (IgG) specific secondary antibodies and photographed at 200x magnification using fluorescent microscopy with the correct excitation and emission filters. [N.B. Since these assays resulted in the formation of large *Leptospira* spp. aggregates, rather than discrete slide-bonded bacteria, the results were clearly either positive (+) or negative (−) at this low 200x magnification and therefore no size bars are shown]

In addition, the S2 serum samples from Group A (DENV-*Leptospira* spp. co-seropositive) patient A.4 and A.5 S2 serum samples, which were positive in *Leptospira* spp. MATs **(Table 1)**, were only IgG-positive in these IFAs **(Figure 2)** while the S2 samples from patient A.6 was both IgM- and IgG-positive in these *Leptospira* spp. IFAs, despite their S1 samples being negative in the *LipL32* gene PCR and their S2 samples, and also being negative in the commercial anti-*Leptospira* spp. IgM ELISA **(Table 1)**. As expected, the S2 serum samples from Group B (DENV seropositive only: i.e. anti-*Leptospira* spp. negative control) patient B.2 was IgM- and IgG-negative in the IFAs **(Figure 2)**, as it also was in the *Leptospira* spp. MAT, and the commercial anti- *Leptospira* spp. IgM ELISA **(Table 1)**, while Group C (*Leptospira* spp. seropositive only: i.e. anti-*Leptospira* spp. positive control) patient C.2 was, as expected, IgM- and IgG- positive in these IFAs **(Figure 2)** as it was in both the *Leptospira* spp. MAT, and the indirect anti-*Leptospira* spp. IgM ELISA **(Table 1)**.

As such, there appeared to be similar results between those obtained in the anti-DENV IgM- and IgG- capture ELISAs and these IgM- and IgG- anti-*Leptospira interrogans* IFAs using the serum samples from Group A (DENV-*Leptospira* spp. co-seropositive) patient A.1 to A.6 **(Table 1, Figure 1 and 2)**, thereby suggesting that these patients’ antibodies could define common epitopes on the outer surface proteins of DENV and pathogenic and non-pathogenic *Leptospira* spp., and which was further investigated.

### Patients’ IgM and IgG titers against paraformaldehyde-fixed *Leptospira* spp. of different serovars in immunofluorescent assays

We further investigated whether there was a correlation between the Group A (DENV-*Leptospira* spp. co- seropositive) A.1 to A.6 IgM/IgG S2 serum sample reaction patterns against the DENVs in the IgM- and IgG- capture ELISAs **(Table 1)** and their IgM/IgG reaction patterns in IFAs against paraformaldehyde-fixed samples of either: i) the non-pathogenic *Leptospira biflexa* Patoc serovar, and ii) a panel of pathogenic *Leptospira interrogans* and intermediately pathogenic *Leptospira fainei* Hurstbridge serovars **(Table 2)**. In this study, the S2 serum sample IgM/IgG positive/negative (+/−) reaction patterns of Group A patient A.1 to A.6 S2 showed the same reaction patterns against the non-pathogenic, pathogenic and intermediately pathogenic *Leptospira* spp. serovars **(Table 2)** as was obtained with them in the anti-DENV IgM- and IgG- capture ELISAs **(Table 1)**. As such, the S2 serum samples from Group A patient A.1 to A.6 showed +/+, +/+, +/−, −/+, −/+ and +/+ IgM/IgG reaction patterns, respectively **(Table 1 and 2)**. In contrast, the S2 samples from Group B (DENV seropositive only: i.e. anti-*Leptospira* spp. negative control) patient B.1 to B.3 all, as expected, showed a −/− IgM/IgG reaction pattern in the IFAs against any of the *Leptospira* spp. serovars **(Table 2)**, despite showing a +/+, −/+ and +/+ IgM/IgG reaction pattern in the anti-DENV IgM-/IgG- capture ELISAs **(Table 1)** and the S2 samples from Group C (*Leptospira* seropositive only: i.e. anti-*Leptospira* positive control) patient C.1 to C.3, as expected, all showed a +/+ IgM/IgG reaction pattern in the IFAs against these different *Leptospira* spp. serovars **(Table 2)**, while they were all negative (−/−) in the anti-DENV IgM- and/or IgG- capture ELISAs **(Table 1)**.

**Table 2.**
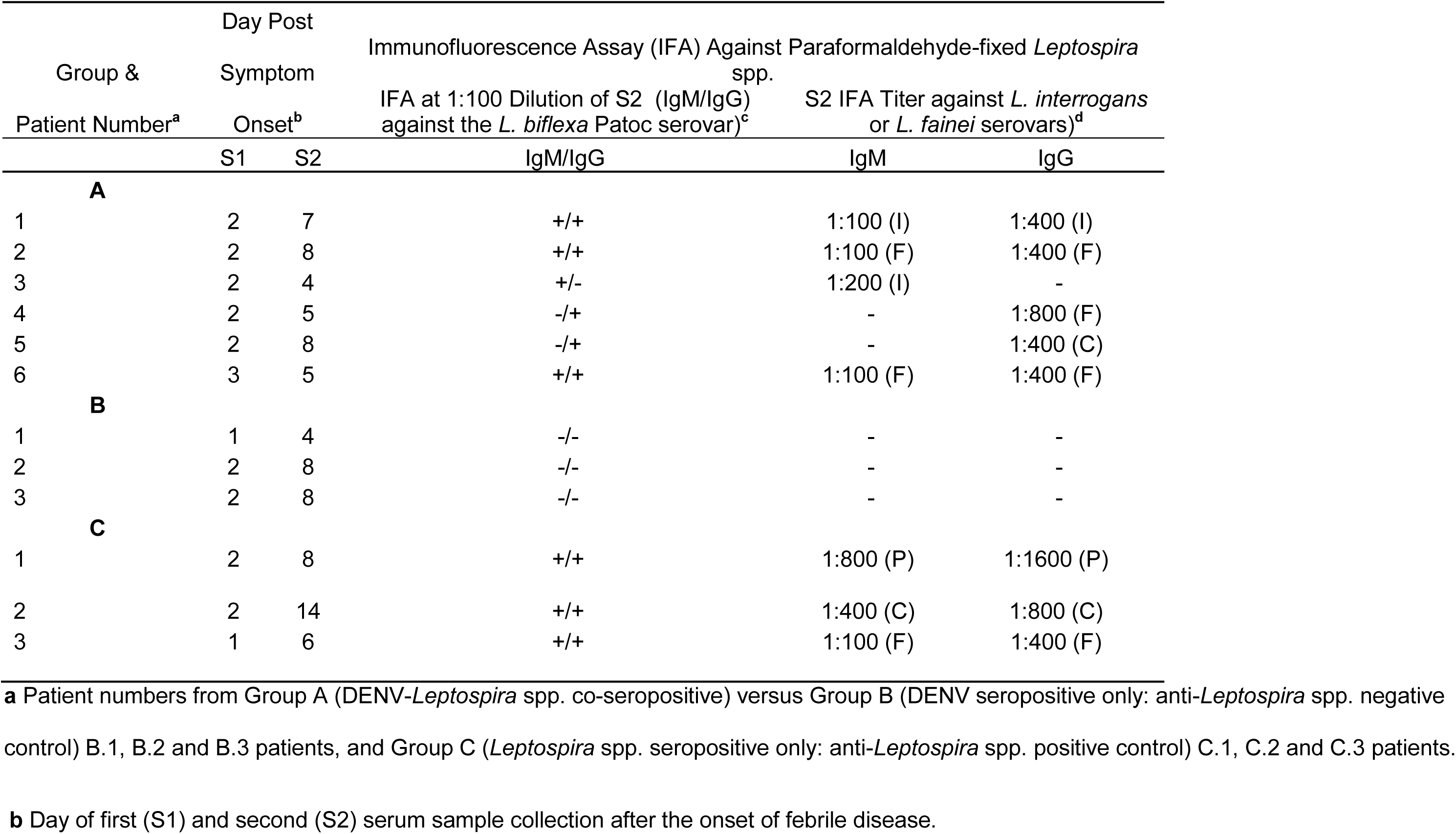
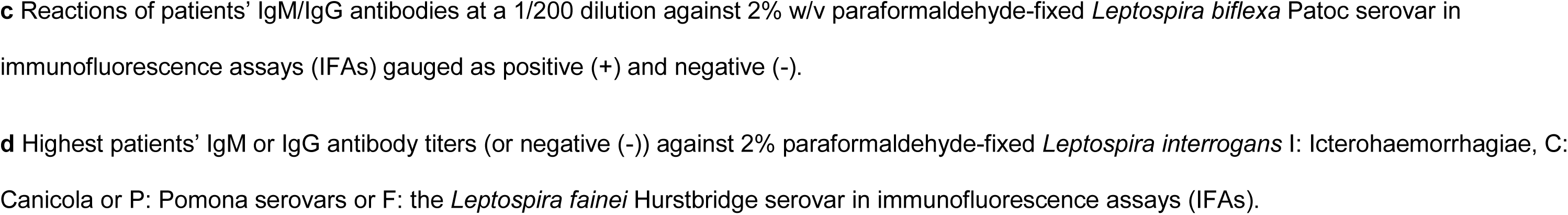
Serological reactions of Group A (DENV-*Leptospira* spp. co-seropositive) versus Group B (DENV seropositive only) and Group C (Leptospira spp. seropositive only) patients in *Leptospira* spp. immunofluorescence assays (IFAs) against members of the *Leptospira interrogans*, *biflexa* or *fainei serovars*.

| Group &<br>Patient Number<sup>a</sup> | Day Post<br>Symptom |  | Immunofluorescence Assay (IFA) Against Paraformaldehyde-fixed *Leptospira* spp. |  |  |
| --- | --- | --- | --- | --- | --- |
|  | Onset<sup>b</sup> |  | IFA at 1:100 Dilution of S2 (IgM/IgG) against the *L. biflexa* Patoc serovar)<sup>c</sup> | S2 IFA Titer against *L. interrogans* or *L. fainei* serovars)<sup>d</sup> |  |
|  | S1 | S2 | IgM/IgG | IgM | IgG |
| **A** |  |  |  |  |  |
| 1 | 2 | 7 | +/+ | 1:100 (I) | 1:400 (I) |
| 2 | 2 | 8 | +/+ | 1:100 (F) | 1:400 (F) |
| 3 | 2 | 4 | +/- | 1:200 (I) | - |
| 4 | 2 | 5 | -/+ | - | 1:800 (F) |
| 5 | 2 | 8 | -/+ | - | 1:400 (C) |
| 6 | 3 | 5 | +/+ | 1:100 (F) | 1:400 (F) |
| **B** |  |  |  |  |  |
| 1 | 1 | 4 | -/- | - | - |
| 2 | 2 | 8 | -/- | - | - |
| 3 | 2 | 8 | -/- | - | - |
| **C** |  |  |  |  |  |
| 1 | 2 | 8 | +/+ | 1:800 (P) | 1:1600 (P) |
| 2 | 2 | 14 | +/+ | 1:400 (C) | 1:800 (C) |
| 3 | 1 | 6 | +/+ | 1:100 (F) | 1:400 (F) |

**a** Patient numbers from Group A (DENV-*Leptospira* spp. co-seropositive) versus Group B (DENV seropositive only: anti-*Leptospira* spp. negative control) B.1, B.2 and B.3 patients, and Group C (*Leptospira* spp. seropositive only: anti-*Leptospira* spp. positive control) C.1, C.2 and C.3 patients.
**b** Day of first (S1) and second (S2) serum sample collection after the onset of febrile disease.
**c** Reactions of patients' IgM/IgG antibodies at a 1/200 dilution against 2% w/v paraformaldehyde-fixed *Leptospira biflexa* Patoc serovar in immunofluorescence assays (IFAs) gauged as positive (+) and negative (-).
**d** Highest patients' IgM or IgG antibody titers (or negative (-)) against 2% paraformaldehyde-fixed *Leptospira interrogans* I: Icterohaemorrhagiae, C: Canicola or P: Pomona serovars or F: the *Leptospira fainei* Hurstbridge serovar in immunofluorescence assays (IFAs).

As such, there was a direct correlation between the IgM- and/or IgG- S2 serum sample reactions of Group A (DENV-*Leptospira* spp. co-seropositive) patient A.1 to A.6 in both the anti-DENV IgM- and/or IgG- capture ELISA reactions and those obtained in these IFAs against these non-pathogenic, pathogenic, and intermediately pathogenic *Leptospira* spp. serovars.

We then further investigated whether there was a correlation between the highest titers obtained using the S2 serum samples of Group A (DENV-*Leptospira* spp. co-seropositive) patient A.1 to A.6 against a particular *Leptospira* spp. serovar in: a) the MATs **(Table 1)** compared to b) the IFAs **(Table 2)**. In this study, the maximum S2 serum sample titers of: i) 1:1600 **(Table 1)** and 1:100 (IgM)/1:400 (IgG) **(Table 2)** were obtained against the *Leptospira interrogans* Icterrohaemorrhagic serovar by patient A.1, ii) 1:3200 **(Table 1)** and 1:100 (IgM)/1:400 (IgG) **(Table 2)** were obtained against the *Leptospira fainei* Hurstbridge serovar by patient A.2, iii) 1:400 **(Table 1),** and 1:200 (IgM)/< 1:100 (IgG) **(Table 2)** were obtained against the *Leptospira interrogans* Icterrohaemorrhagic serovar with patient A.3, iv) 1:400 **(Table 1)** and < 1:100 (IgM)/1:800 (IgG) **(Table 2)** were obtained against *Leptospira fai*nei Hurstbridge serovar by patient A.4, v) 1:800 **(Table 1)** and < 1:100 (IgM)/1:400 (IgG) **(Table 2)** were obtained against the *Leptospira interrogans* Canicola serovar with patient A.5, and vi) 1:400 **(Table 1)** and 1:100 (IgM)/1:400 (IgG) **(Table 2)** were obtained against *Leptospira fainei* Hurstbridge serovar by patient A.6. As such, while slightly lower titers were obtained in the IgM- and/or IgG-specific IFAs **(Table 1)** than in the MATs which detected combined IgM/IgG titers **(Table 2),** these assays showed the same reaction patterns against a particular *Leptospira* spp. serovar where the highest IFA titers amongst the Group A patients were obtained using their S2 serum samples against the intermediately pathogenic *Leptospira fainei* Hurstbridge serovar (n = 3: patient A.2, A.4 and A.6), followed by the highly pathogenic *Leptospira interrogans* Icterrohaemorrhagiae serovar (n = 2: patient A.1 and A.3) and the pathogenic *Leptospira interrogans* Canicola serovar (n = 1: patient A.5).

As expected, the S2 serum samples from Group B (DENV seropositive only: i.e. anti-*Leptospira* spp. negative control) patient B.1 to B.3 showed no reactions against *Leptospira* spp. in either the MATs or IFAs, while those of Group C (*Leptospira* spp. seropositive only: i.e. anti-*Leptospira* spp. positive control) patient C.1 to C.3 also showed a correlation between the maximum MAT and IFA titers as: i) 1:3200 **(Table 1)** and 1:800 (IgM)/1600 (IgG) **(Table 2)** being obtained against the pathogenic *Leptospira interrogans* Pomona serovar by patient C.1, ii) 1:3200 **(Table 1)** and 1:400 (IgM)/1:800 (IgG) **(Table 2)** titers being obtained against the *Leptospira interrogans* Canicola serovar by patient C.2, and iii) 1:400 **(Table 1)** and 1:100 (IgM)/1:400 (IgG) **(Table 2)** titers were obtained against the *Leptospira fainei* Hurstbridge serovar by patient C.3.

### Protein (Western) blotting

Protein (Western) blotting was then used to identify the target protein antigen types identified by these IgM and/or IgG antibodies on different *Leptospira* spp. serovars. In this study, very high numbers (1 x 10^9^/ml) of the *Leptospira interogans* Icterohaemorhagiae (ICT), Tarassovi (TAR) and Canicola (CAN), *Leptospira fainei* (FAI) Hurstbridge, and the *Leptospira biflexa* Patoc (PAT) serovars were treated with lysis buffer and either left non-reduced or were reduced with 2-mercaptoethanol (2-ME) for the initial antigenic comparisons **(Figure 3)**. In this study, the S2 serum sample from Group C (*Leptospira* spp. seropositive only: i.e. anti-*Leptospira* spp. positive control) patient C.1 clearly showed reactions with major protein bands of approximately 68-72 and 38-42 KDa of each of these pathogenic, intermediately pathogenic and non-pathogenic *Leptospira* spp. serovars, and which showed slightly increased reactions when these samples were reduced by heating with 2- mercaptoethanol (2-ME). These two bands of the *Leptospira interrogans* Canicola (CAN) and the *Leptospira biflexa* Patoc (PAT) serovars, however, also displayed some possible truncations detected as approximately 56- 60 and 34-36 KDa bands.

**Figure 3.**
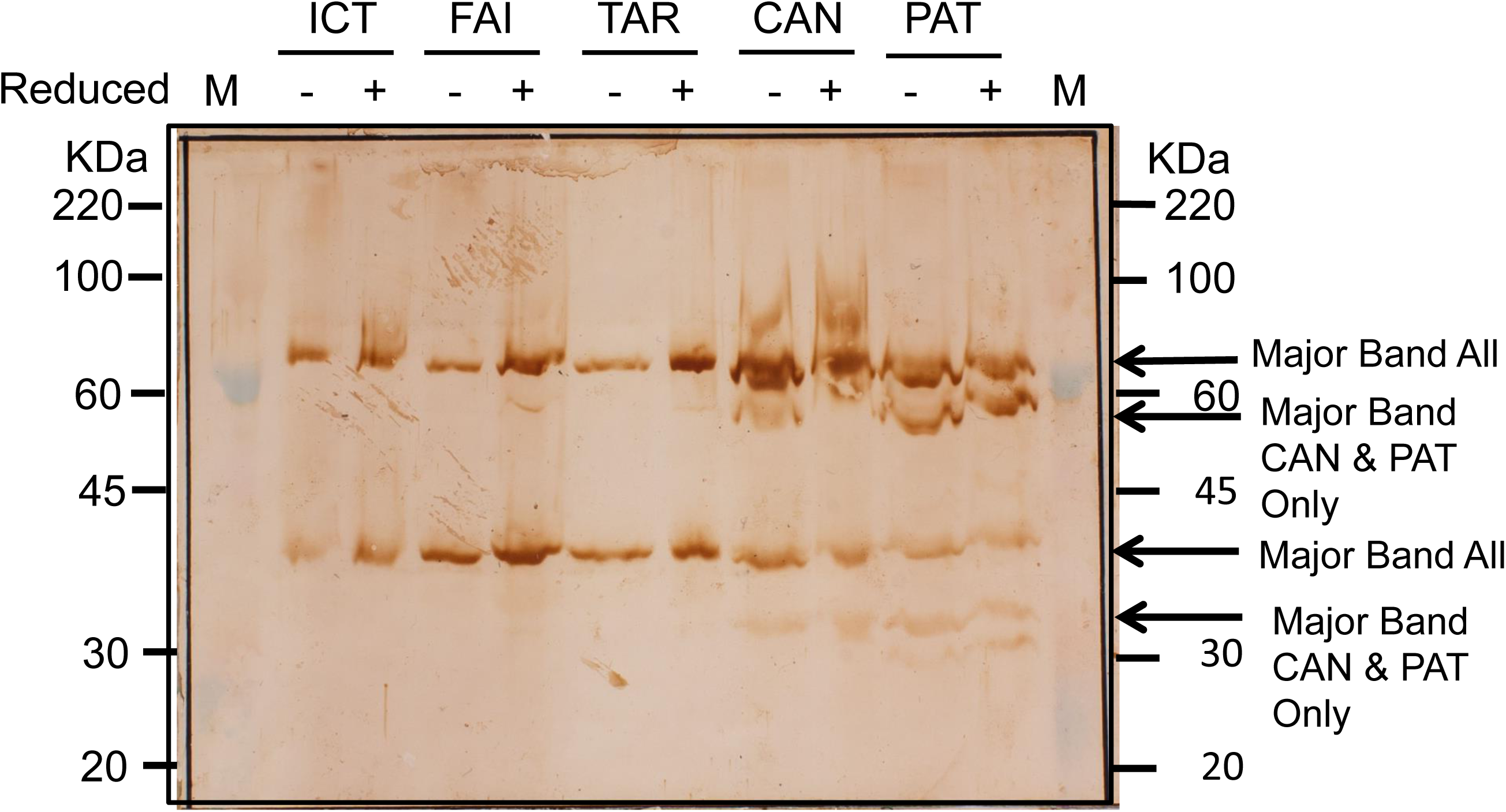
Immunoblot of *Leptospira* spp. cell lysates of different serovars using *Leptospira* spp. reactive human sera. Washed *Leptospira interrogans* of the Icterrohaemorrhagiae (ICT), Tarrasovi (TAR) and Canicola (CAN) seovars, the *Leptospira biflexa* serovar Patoc (PAT) serovar and the *Leptospira fainei* (FAI) Hurstbridge serovar were heated in SDS lysis buffer with (+) or without (−) reduction by 2-mercaptoethanol (2-ME) and the lysates of approximately 1-2 x 10^8^ bacteria were loaded into each lane. The S2 serum sample of Group C (*Leptospira* spp. seropositive only: i.e. anti-*Leptospira* spp. positive control) patient C.1 was diluted at 1/200 and reacted with the blot and the bound antibodies were detected by sequential reaction steps with peroxidase-labelled anti-human IgG (H&L) secondary antibodies and DAB/4C1N substrate. The 20-220 KDa rainbow molecular weight markers are shown and the dominant *Leptospira* spp. bands of approximately 68-72 KDa and 38-42 KDa are shown, but bands of approximately 56-60 and 34-36 KDa were also detected in the lysates of the CAN and PAT serovars.

These same 68-72 and 38-42 KDa molecular weight bands were also observed in the non-reduced *Leptospira interrogans* Icterohaemorrhagiae serovar lysate when reacted with the S2 serum samples from Group A (DENV-*Leptospira* spp. co-seropositive) patient A.1 to A.4 (immunoblot strips 1-4), respectively (Figure 4), but those of patient A.1 and A.3 displayed stronger reactions against the 68-72 KDa band while the samples of patient A.2 and A.4 displayed stronger reactions with the 38-42 KDa protein band, while the S2 serum sample mixture from patient A.5 and A.6 (strip 7) showed weaker reactions against both these 68-72 and 38-42 KDa bands but a much stronger reaction against an 18 KDa protein band **(Figure 4)**. The S2 serum sample from Group C (*Leptospira* spp. seropositive only: i.e. anti-*Leptospira spp.* positive control) patient C.1 (strip 5) also showed similar but much stronger reactions against both the 68-72 and 38-42 KDa protein bands, while no reactions were observed with the S2 serum sample of Group B (DENV seropositive only: anti-*Leptospira* spp. positive control) patient B.1 (strip 6) with any of these protein bands (strip 6). The reactions with an approximately 18 KDa protein was also observed using the S2 serum samples of Group A patient A.1-A.4 and Group C patient C.1, but not with that of Group B (DENV seropositive only: anti-*Leptospira* spp. negative control) patient B.1. As such, this 18 KDa protein band, which was very strongly identified by the S2 serum sample mixture of patients A.5/A.6 (strip 7), but which less strongly reacted with the 68-72 and 38-42 KDa bands therefore suggested that it was likely to be another truncated product of these higher molecular weight proteins due to further protease activity. This was supported by the observations that this reaction with this 18 KDa band was not observed in the earlier immunoblot reactions of the S2 serum sample of Group C (*Leptospira* spp. seropositive only) patient C.1 with the same *Leptospira interrogans* Icterrohaemorrhagiae serovar **(Figure 3)**. Interestingly, these sera also all showed reactions with two high molecular weight protein bands of approximately 300 and 260 KDa, either suggesting that they: a) were protein aggregates of either the *Leptospira interrogans* Icterohaemorrhagiae serovar or b) were non-specific reactions due to possible cross-reactions between the labelled secondary antibodies with other non-*Leptospira* spp. proteins present in the media of these *Leptospira* spp. lysates, which could possibly account for the reaction even using the S2 serum sample from Group B (DENV seropositive only: i.e. *Leptospira* spp. negative control) patient B.1 **(Figure 4: strip 6)**. To further test these possibilities, the reactions of non-reduced and 2-ME reduced *Leptospira interrogans* Icterohaemorrhagiae (ICT) and *Leptospira biflexa* Patoc (PAT) serovar lysates were compared using the S2 serum samples from Group C (*Leptospira* spp. seropositive only: i.e. anti-Leptospira positive control) patient C.1 (strips 1 and 4) with that of Group A patient A.1 (DENV- *Leptospira* spp. cross-reactive: secondary DENV infection) (strips 2 and 5) and patient A.3 (DENV and *Leptospira* spp. cross-reactive: primary DENV infection) (strips 3 and 6) **(Figure 5)**. In this study, all three of these S2 sera displayed strong reactions with the approximate Mr bands of approximately 300 and 260 KDa and the 68-72 and 38-42 KDa protein bands, as well as their likely truncated 34-36 and 18 KDa protein bands in the non-reduced lysates of these different *Leptospira* spp. serovars. In contrast, the reactions with these approximately 300 and 260 KDa major bands was abrogated, while their reactions with the 68-72 and 38-42 KDa bands were increased, particularly with those of the *Leptospira interrogans* Icterrohaemorrhagiae serovar, when these lysates were reduced. As such, these results suggested that reduction with 2-ME resulted in the dissociation of these high 200 and 260 KDa molecular weight aggregates of the 68-72 and 38-42 KDa proteins in these lysates, and the apparent reaction of the S2 serum sample of Group B (DENV seropositive only: i.e. anti-Leptospira spp. negative control) with these 300 and 260 KDa aggregates, but not the 68-72, 38-42 *Leptospira* spp. proteins, could have occurred due to non-specific binding of either: a) their IgM or IgG antibodies, or b) the peroxidase-labelled goat anti-human IgG (H&L) secondary polyclonal antibodies **(Figure 4: strip 6)**.

**Figure 4.**
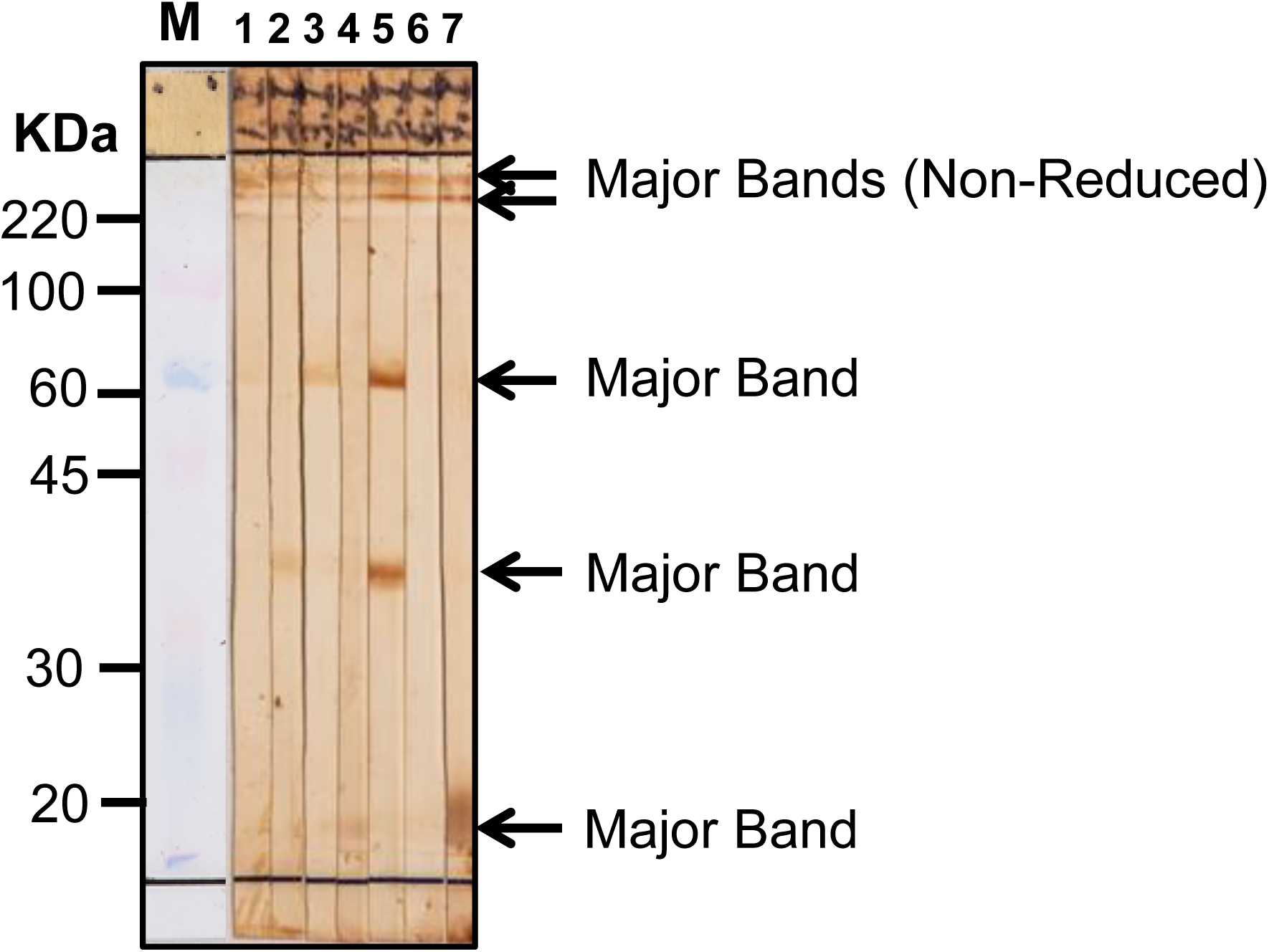
Immunoblot of *Leptospira* spp. cell lysates with different patients’ serum samples. Washed *Leptospira interrogans* of the highly pathogenic Icterrohaemorrhagiae (ICT) serovar were lysed with non-reducing SDS lysis buffer and preparative strips were cut and reacted with a 1/200 dilution of the S2 serum samples from Group A (DENV and *Leptospira* spp. co-seropositive) patient A.1 (strip 1), A.2 (strip 2), A.3 (strip 3), A.4 (strip 4) and a A.5/.6 mixture (strip 7), while those from Group C (*Leptospira* spp. seropositive only: anti-*Leptospira* spp. positive control) patient C.1 (strip 5), and Group B (DENV seropositive only: i.e. anti- *Leptospira* spp. negative control) patient B.1 (strip 6). Their bound antibodies were subsequently detected by sequential reaction steps with peroxidase-labelled anti-human IgG (H&L) secondary antibodies and DAB/4C1N substrate. The 20-220 KDa rainbow molecular weight markers are shown and the major *Leptospira* spp. bands of approximately 300, 260, 68-72, 38-42 and 18 KDa are shown.

**Figure 5.**
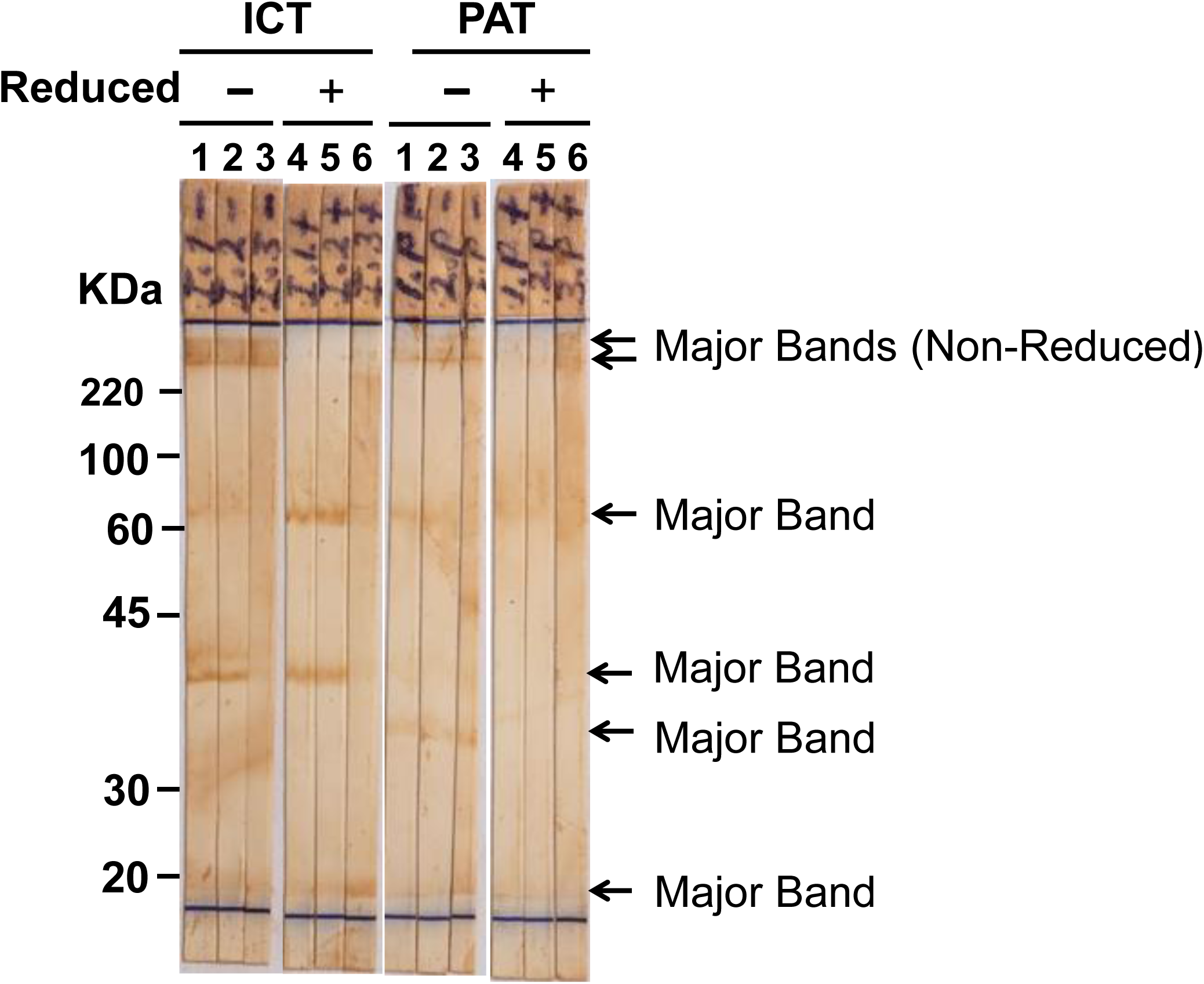
Immunoblot of pathogenic and non-pathogenic *Leptospira* spp. serovars with different patients’ serum samples. Washed *Leptospira interrogans* of the highly pathogenic Icterohaemorrhagiae (ICT) or non-pathogenic *Leptospira biflexa* Patoc (PAT) serovars were either heated with reducing (+) or non-reducing (−) SDS lysis buffer, and subjected to preparative immunoblotting using 1/200 dilutions of the S2 serum samples from Group C (*Leptospira* spp. seropositive only: i.e. *Leptospira* spp. positive control) patient C.1 (strips 1 and 4) or from two patients from Group A (DENV and *Leptospira* spp. co-seropositive) which had either on-going secondary (patient A.1: strips 2 and 5) or primary (patient A.3: strips 3 and 6) DENV infections with non-reduced (strips 1- 3) or reduced (strips 4-5) lysates of the Icterohaemorrhagiae (ICT) or Patoc (PAT) serovars, and their bound antibodies were detected by sequential reaction steps with peroxidase-labelled anti-human IgG and IgM (H&L) secondary antibodies and DAB/4C1N substrate. The 20-220 KDa rainbow molecular weight markers are shown and the major *Leptospira* spp. bands detected by each of the S2 serum samples from patient A.1, A.3 and C.1 of approximately 300, 260, 68-72, 38-42 and 18 KDa are shown by bold black arrows, while only the reactions with the high molecular weight 300 and 260 KDa bands were abrogated by reduction with 2ME. The major truncated band of approximately 34-36 KDa was particularly visible in the non-reduced lysate of the PAT serovar.

### Clinical details of Group A, B and C patients

Clinically-graded details of these Group A patients did not show the classical features of leptospirosis (e.g.conjunctival suffusion, etc.), as were displayed by the Group C patients **(3)** and therefore these Group A (DENV-*Leptospira* spp. co-seropositive) patients only displayed the symptoms of classical dengue fever (DF) as did the Group B (DENV seropositive only) patients **(5,6)**, and none of our 70 patients with confirmed on- going DENV infections, who subsequently developed severe dengue (dengue hemorrhagic fever/dengue shock syndrome) **(5,6)** were positive in the *Leptospira* spp. MATs performed in this study. As such, these finding further supported the belief that these Group A patients only had on-going DENV infections, but who also generated cross-reactive IgM and/or IgG antibodies to *Leptospira* spp. outer surface membrane antigens at such elevated titers to cause false-positive reactions in the ‘gold standard’ MAT and the IFAs, rather than being true DENV-*Leptospira* spp. co-infections, which have been reported previously (see Discussion).

## Discussion

We believe that this was the first description of antibody cross-reactions of DENV primary (IgM only) or secondary (IgG with or without IgM) patients’ antibodies with outer surface proteins of all *Leptospira* spp. serovars tested in the ‘gold standard’ *Leptospira* spp. MAT, IFAs and immunoblot (western blot) assays, thereby resulting in strong false positive reactions. These results were clearly shown in our study by the rising S1 to S2 serum samples titers against both DENV and *Leptospira* spp. due to the collection and testing of paired serum samples, but which is not routinely performed by laboratories throughout the world.

Since the antibody reactions in the DENV IgM- and IgG- capture ELISA are predominantly generated against the major, immunodominant envelope (E) glycoprotein (E gp) present on DENV virions, common linear (sequential) epitope sequences on the DENV E gp. and *Leptospira* spp. 68-72 and 38-42 KDa proteins need to be identified since these cross-reactions with the *Leptospira* spp. antigens were strongly identified even in their 2-ME reduced lysates. Many sequential (linear) epitopes have been identified on the DENV E gps. **(e.g. 20, 22- 26)**, but since only a few (8.6%: 6/70) of our patients with confirmed DENV primary or secondary infections were able to cross-react with the *Leptospira* spp. outer surface membrane antigens on all serovars tested, immunodominant epitopes on the DENV E gps, such as the partially cryptic fusion sequence (101- WGNGCGLF-109) **(22–25)** and others such as the ELK/KLE and SxK/KxS type motifs **(27, 28)** are unlikely suspects. Further, since Group A contained one case of a primary DENV infection from which DENV-2 was isolated (patient A.3) , common linear, sequential B-cell epitopes on the immuno-dominant DENV-2 E gp and *Leptospira* spp. outer surface membrane proteins in the 68-72 and 38-42 KDa molecular weight range should be investigated and confirmed using DENV-2 E gp. synthetic peptide sequences and likely epitope sequences on these *Leptospira* spp. outer surface membrane proteins using Group A and C patients’ S2 serum samples.

There have been multiple reports aimed to differentiate DENV and *Leptospira* spp. infections and to also identify co-infections by these pathogens **(7–16)**. In several of these studies however only the ‘gold standard’ *Leptospira* spp. MATs **(9,15,16)** or anti-*Leptospira* spp. IgM and IgG reactions in IFAs **(7)** were used for the diagnosis of the *Leptospira* spp. infections. As such, at least some of these patients with these apparent DENV- *Leptospira* spp. co-infections may have resulted from false positive reactions as we found in our study. This possibility should therefore be further tested using either isolation in complete EMJH medium or using a *Leptospira* spp. specific PCR with their S1 serum samples and anti-*Leptospira* spp. ELISAs or lateral flow assays, preferably using their paired S1 and S2 serum samples to confirm or exclude that these patients also had ‘on-going’ *Leptospira* spp. infections as was tested in our study.

While the ‘gold standard’ *Leptospira* spp. MAT is highly specific and detects total immunoglobulins, it has been reported to be less sensitive than the commercial IgM latex agglutination assay, but was equally sensitive to an anti-*Leptospira* spp. IgM ELISA in one study **(13).** There have however been different reports on the suitability of anti-*Leptospira* spp. IgM ELISAs versus the MATs and while many studies promoted the use of the anti-*Leptospira* spp. IgM ELISAs versus the MAT for very early acute phase diagnosis **(e.g. 29,30)**, as was recently reviewed **(31)**, at least one (e.g. the commercial Panbio anti-*Leptospira* IgM ELISA) as was reported to result in very low specificity (60.9%) and sensitivity (65.6%) compared to the MATs in a well-conducted comparison **(32)**. In addition, the use of only few or even single antigens in these different commercial anti- *Leptospira* spp. IgM ELISAs and lateral flow assays was stated to be a distinct disadvantage of them by the WHO **(3)**. The negative results using the commercial Standard Diagnostics Inc. anti-*Leptospira* spp. IgM ELISA in our study, despite positivity in the MAT and IgM- and IgG- anti-*Leptospira* spp. IFA, strongly suggested that the *Leptospira* spp. antigens bound in the MATs, IFAs and immunoblot (western blot) assays were not present in that commercial ELISA and therefore it did not contain the 68-72 and 38-42 KDa outer surface membrane proteins identified by Group A and C patients’ sera on all of the pathogenic, as well as non-pathogenic, *Leptospira* spp. serovars tested. The *Leptospira biflexa* Patoc serovar (non-pathogenic ‘saprophytic’ strain) and the *Leptospira fainei* Hurstbridge serovar (‘intermediately’ pathogenic isolate) used in our study were previously shown to be negative in *LipL32* PCR assays **(19)**, but 40% (2/5) of the Group C (*Leptospira* spp. seropositive only) patients’ S2 sera showed the highest MAT titres against this *Leptospira* spp. serovar but which were also *LipL32* PCR positive **(Table 1)**. While the *Leptospira fainei* Hurstbridge serovar can cause serious Weil’s-like disease in humans **(33)** and the highest MAT titers against a particular *Leptospira* spp. serovar in MATs are indicative, there are many reports that they are not reliable in identifying the particular *Leptospira* spp. infecting serovar **(3,34,35)**. As such, the actual infecting *Leptospira* spp. serovars identified using our patients’ S2 serum samples, including the *Leptospira fainei* Hurstbridge serovar **(Table 1 and 2)** may not have been correctly identified by the Group A and C patients’ sera in the MATs and IFAs.

While many studies have been performed to identify outer surface membrane proteins specific to pathogenic *Leptospira interrogans* for diagnostic, pathogenesis and vaccine studies **(31,36,37)**, the 68-72 and 38-42 KDa outer surface membrane proteins found in our study were common to pathogenic *Leptospira interrogans*, the intermediately pathogenic *Leptospira fainei* Hurstbridge, as well as the non-pathogenic *Leptospira biflexa* Patoc serovars **(Figure 1)** this non-pathogenic serovar was found to possess approximately 1/3 less protein-encoding genes that pathogenic *Leptospira interrogans* serovars **(38)**. Thus, proteomic analyses will be required to specifically identify these common 68-72 and 38-42 KDa *Leptospira* spp. outer surface membrane proteins.

Importantly, such early acute-phase diagnosis of leptospirosis is vital to provide these patients with prompt antibiotic therapy such as early empirical doxycycline treatment, which was the most efficient and cost-effective regimen to shorten the duration of fever and prevent patient complications **(39)**, and which otherwise could result in a severe leptospirosis death rate of up to 50% even with intensive care unit (ICU) support within modern facilities **(40)**. Our Group A (DENV-*Leptospira* spp. co-seropositive) A.1 to A.6 patients did not show clinically graded differential DENV or leptospirosis diagnostic criteria **(6)**. However, without assessing whether or not patients have true DENV-*Leptospira* co-infections using the appropriate differential diagnostic assays and simple differential clinical and prognostic criteria **(6)**, some patients may not achieve the essential early antibiotic therapy for leptospirosis or the early hospital-based supportive therapy required to avert/block or lessen the subsequently development of life-threatening severe dengue (DHF/DSS: dengue hemorrhagic fever/dengue shock syndrome) **(6)**.

## Data Availability

All data produced in the present work are contained in the manuscript

## Acknowledgements

This study was performed under Regalias Desarrollo de un Programa de CT+I en Enfermedades Infecciosas Todo El Departamento, Atlántico, Caribe research grant number from 0103-2015-000039 entitled ‘Implementación de métodos serológicos, moleculares para la caracterización de las fiebres hemorrágicas producidas por virus dengue y la bacteria *Leptospira* spp. en humanos en Soledad y Distrito de Barranquilla durante los años de 2014-2016’. We also thank Alvaro Cepeda Samudio from the Archivo Audiovisual (Universidad del Norte) for preparing the high-resolution photographs of the immunoblots for Figure 3-5.

## Notes

### Competing Interest Statement

The authors have declared no competing interest.

### Funding Statement

Regalias Desarrollo de un Programa de CT+I en Enfermedades Infecciosas Todo El Departamento, Atlantico, Caribe research grant number from 0103-2015-000039

### Author Declarations

The Ethics Committee from Universidad del Norte gave ethical approval for this work - All necessary patient/participant consent has been obtained - Any patient/participant/sample identifiers cannot be used to identify individuals

## References

1. World Health Organization1997. Dengue haemorrhagic fever: diagnosis, treatment, prevention and control, 2nd ed. World Health Organization Geneva, Switzerland https://apps.who.int/iris/handle/10665/41988

2. World Health Organization 2009). Dengue guidelines for diagnosis, treatment, prevention and control: new edition. World Health Organization Geneva, Switzerland. https://apps.who.int/iris/handle/10665/44188

3. World Health Organization 2003. Human leptospirosis: guidance for diagnosis, surveillance and control. World Health Organization Geneva, Switzerland https://apps.who.int/iris/handle/10665/42667

4. Costa F, Hagan JE, Calcagno J, Kane M, Torgerson P, Martinez-Silveira MS, Stein C, Abela-Ridder B, Ko AI. 2015. Global Morbidity and Mortality of Leptospirosis: A Systematic Review. PLoS Negl Trop Dis 9:e0003898. https://doi10.1371/journal.pntd.0003898.

5. Falconar AK, de Plata E, Romero-Vivas CM. 2006. Altered enzyme-linked immunosorbent assay immunoglobulin M (IgM)/IgG optical density ratios can correctly classify all primary or secondary dengue virus infections 1 day after the onset of symptoms, when all the viruses can be isolated. Clin Vaccine Immunol 13:1044–1051. https://doi10.1128/CVI.00105-06.

6. Falconar AK, Romero-Vivas CM. 2012. Simple Prognostic Criteria Can Definitively Identify Patients who Develop Severe Versus Non-Severe Dengue Disease, or Have Other Febrile Illnesses. J Clin Med Res 4:33–44. https://doi10.4021/jocmr694w.

7. Arroyave E, Londoño AF, Quintero JC, Agudelo-Flórez P, Arboleda M, Díaz FJ, Rodas JD. 2013. Etiology and epidemiological characterization of non-malarial febrile syndrome in three municipalities of Urabá (Antioquia), Colombia. Biomedica 33 Suppl 1:99–107. PMID: 24652254.

8. Chaudhry R, Das A, Premlatha MM, Choudhary A, Chourasia BK, Chandel DS, Dey AB. 2013. Serological & molecular approaches for diagnosis of leptospirosis in a tertiary care hospital in north India: a 10-year study. Indian J Med Res 2013 137:785–790. PMID: 23703348.

9. Rodríguez-Villamarín FR, Edgar Prieto-Suárez E, Patricia L Escandón PL, de la Hoz Restrepo F. 2014. Leptospirosis percentage and related factors in patients having a presumptive diagnosis of dengue, 2010-2012. Rev Salud Publica (Bogota) 16:597–609. PMID: 25791310

10. Pérez Rodríguez NM, Galloway R, Blau DM, Traxler R, Bhatnagar J, Zaki SR, Rivera A, Torres JV, Noyd D, Santiago-Albizu XE, Rivera García B, Tomashek KM, Bower WA, Sharp TM. 2014. Case series of fatal Leptospira spp./dengue virus co-infections-Puerto Rico, 2010-2012. Am J Trop Med Hyg 91:760–765. https://doi:10.4269/ajtmh.14-0220.

11. de Melo Bezerra LF, Fontes RM, Gomes AM, da Silva DA, Colares JK, Lima DM. 2015. Serological evidence of leptospirosis in patients with a clinical suspicion of dengue in the State of Ceará, Brazil. Biomedica 35:557–562. https://doi10.7705/biomedica.v35i4.2504.

12. Mørch K, Manoharan A, Chandy S, Chacko N, Alvarez-Uria G, Patil S, Henry A, Nesaraj J, Kuriakose C, Singh A, Kurian S, Gill Haanshuus C, Langeland N, Blomberg B, Vasanthan Antony G, Mathai D. 2017. Acute undifferentiated fever in India: a multicentre study of aetiology and diagnostic accuracy. BMC Infect Dis. 17:665. https://doi10.1186/s12879-017-2764-3.

13. Suppiah J, Chan SY, Ng MW, Khaw YS, Ching SM, Mat-Nor LA, Ahmad-Najimudin NA, Chee HY. 2017. Clinical predictors of dengue fever co-infected with leptospirosis among patients admitted for dengue fever - a pilot study. J Biomed Sci. 24:40. https://doi10.1186/s12929-017-0344-x.

14. Sachu A, Madhavan A, Vasudevan A, Vasudevapanicker J. 2018 Prevalence of dengue and leptospirosis co-infection in a tertiary care hospital in south India. Iran J Microbiol 10:227–232. PMID: 30483374.

15. Cardona-Ospina JA, Jiménez-Canizales CE, Vásquez-Serna H, Garzón-Ramírez JA, Alarcón-Robayo JF, Cerón-Pineda JA, Rodríguez-Morales AJ. 2018. Fatal Dengue, Chikungunya and Leptospirosis: The Importance of Assessing Co-infections in Febrile Patients in Tropical Areas. Trop Med Infect Dis 3:123. https://doi10.3390/tropicalmed3040123.

16. Hishamshah M, Ahmad N, Mohd Ibrahim H, Nur Halim NA, Nawi S, Amran F. 2018. Demographic, clinical and laboratory features of leptospirosis and dengue co-infection in Malaysia. J Med Microbiol 67:806–813. https://doi10.1099/jmm.0.000750.

17. Romero-Vivas CM, Cuello-Pérez M, Agudelo-Flórez P, Thiry D, Levett PN, Falconar AK. 2013a. Cross-sectional study of Leptospira seroprevalence in humans, rats, mice, and dogs in a main tropical sea-port city.Am J Trop Med Hyg. 88:178–183. https://doi10.4269/ajtmh.2012.12-0232.

18. Romero-Vivas CM, Thiry D, Rodríguez V, Calderón A, Arrieta G, Máttar S, Cuello M, Levett PN, Falconar AK. 2013b. Molecular serovar characterization of Leptospira isolates from animals and water in Colombia. Biomedica 33 Suppl 1:179–84. PMID: 24652261.

19. Levett PN, Morey RE, Galloway RL, Turner DE, Steigerwalt AG, Mayer LW. 2005. Detection of pathogenic leptospires by real-time quantitative PCR. J.Med. Microbiol. 54:45–49. doi: 10.1099/jmm.0.45860-0.

20. Falconar AK. 1999. Identification of an epitope on the dengue virus membrane (M) protein defined by cross-protective monoclonal antibodies: design of an improved epitope sequence based on common determinants present in both envelope (E and M) proteins. Arch Virol 144:2313–2330. https://doi10.1007/s007050050646.

21. Pinne M, Haake D. 2011. Immuno-fluorescence assay of Leptospiral surface-exposed proteins. J of Visual Experiment (JoVE) https://doi10.3791/2805.

22. Crill WD, Chang GJ. 2004. Localization and characterization of flavivirus envelope glycoprotein cross-reactive epitopes. J Virol 78:13975–86. https://doi10.1128/JVI.78.24.13975-13986.2004.

23. Stiasny K, Kiermayr S, Holzmann H, Heinz FX. 2006. Cryptic properties of a cluster of dominant flavivirus cross-reactive antigenic sites. J Virol 80:9557–9568. https://doi10.1128/JVI.00080-06.

24. Crill WD, Trainor NB, Chang GJ. 2007. A detailed mutagenesis study of flavivirus cross-reactive epitopes using West Nile virus-like particles. J Gen Virol 88:1169–1174. https://doi10.1099/vir.0.82640-0

25. Falconar AK. 2008a. Monoclonal antibodies that bind to common epitopes on the dengue virus type 2 nonstructural-1 and envelope glycoproteins display weak neutralizing activity and differentiated responses to virulent strains: implications for pathogenesis and vaccines. Clin Vaccine Immunol 15:549–561. https://doi10.1128/CVI.00351-07.

26. Falconar AKI. 2008b. Use of synthetic peptides to represent surface-exposed epitopes defined by neutralizing dengue complex- and flavivirus group-reactive monoclonal antibodies on the native dengue type-2 virus envelope glycoprotein. J Gen Virol 89:1616–1621. https://doi10.1099/vir.0.83648-0.

27. Falconar AK. 1997. The dengue virus nonstructural-1 protein (NS1) generates antibodies to common epitopes on human blood clotting, integrin/adhesin proteins and binds to human endothelial cells: potential implications in haemorrhagic fever pathogenesis. Arch Virol 142:897–916. https://doi10.1007/s007050050127.

28. Falconar AK. 2012. Epitope reactions can be gauged by relative antibody discriminating specificity (RADS) values supported by deletion, substitution and cysteine bridge formation analyses: potential uses in pathogenesis studies. BMC Res Notes. 5:208. https://doi10.1186/1756-0500-5-208.

29. Niloofa R, Fernando N, de Silva NL, Karunanayake L, Wickramasinghe H, Dikmadugoda N, Premawansa G, Wickramasinghe R, de Silva HJ, Premawansa S, Rajapakse S, Handunnetti S. 2015. Diagnosis of Leptospirosis: Comparison between Microscopic Agglutination Test, IgM-ELISA and IgM Rapid Immunochromatography Test. PLoS One 10:e0129236. https://doi10.1371/journal.pone.0129236.

30. Zin NM, Othman SN, Abd Rahman FR, Abdul Rachman AR. 2019. Evaluation of IgM LAT and IgM ELISA as compared to microscopic agglutination test (MAT) for early diagnosis of Leptospira sp. Trop Biomed 36:1071–1080. PMID: 33597476

31. Samrot AV, Sean TC, Bhavya KS, Sahithya CS, Chan-Drasekaran S, Palanisamy R, Robinson ER, Subbiah SK, Mok PL.2021. Leptospiral Infection, Pathogenesis and Its Diagnosis-A Review. Pathogens 10:145. https://doi10.3390/pathogens10020145.

32. Blacksell SD, Smythe L, Phetsouvanh R, Dohnt M, Hartskeerl R, Symonds M, Slack A, Vongsouvath M, Davong V, Lattana O, Phongmany S, Keolouangkot V, White NJ, Day NP, Newton PN. 2006. Limited diagnostic capacities of two commercial assays for the detection of Leptospira immunoglobulin M antibodies in Laos. Clin Vaccine Immunol13:1166–1169. https://doi10.1128/CVI.00219-06.

33. Petersen AM, Boye K., Blom J, Schlichting P, Krogfelt KA. 2001. First isolation of Leptospira fainei serovar Hurstbridge from two human patients with Weil’s syndrome. J Med Microbiol https://doi.org/10.1099/0022-1317-50-1-96

34. Smythe LD, Wuthiekanun V, Chierakul W, Suputtamongkol Y, Tiengrim S, Dohnt MF, Symonds ML, Slack AT, Apiwattanaporn A, Chueasuwanchai S, Day NP, Peacock SJ. 2009. The microscopic agglutination test (MAT) is an unreliable predictor of infecting Leptospira serovar in Thailand. Am J Trop Med Hyg 81:695–697. https://doi:10.4269/ajtmh.2009.09-0252.

35. Murray CK, Gray MR, Mende K, Parker TM, Samir A, Rahman BA, Habashy EE, Hospenthal DR, Pimentel G. 2011. Use of patient-specific Leptospira isolates in the diagnosis of leptospirosis employing microscopic agglutination testing (MAT). Trans R Soc Trop Med Hyg 105:209–213. https://doi:10.1016/j.trstmh.2010.12.004.

36. Haake DA, Zückert WR. 2015. The leptospiral outer membrane. Curr Top Microbiol Immunol 387:187–221. https://doi10.1007/978-3-662-45059-8_8.

37. Raja V, Natarajaseenivasan K. 2015. Pathogenic, diagnostic and vaccine potential of leptospiral outer membrane proteins (OMPs). Crit Rev Microbiol 41:1–17. https://doi10.3109/1040841X.2013.787387

38. Picardeau M, Bulach DM, Bouchier C, Zuerner RL, Zidane N, Wilson PJ, Creno S, Kuczek ES, Bommezzadri S, Davis JC, McGrath A, Johnson MJ, Boursaux-Eude C, Seemann T, Rouy Z, Coppel RL, Rood JI, Lajus A, Davies JK, Médigue C, Adler B. 2008. Genome sequence of the saprophyte Leptospira biflexa provides insights into the evolution of Leptospira and the pathogenesis of leptospirosis. PLoS One. 3:e1607. https://doi10.1371/journal.pone.0001607.

39. Suputtamongkol Y, Pongtavornpinyo W, Lubell Y, Suttinont C, Hoontrakul S, Phimda K, Losuwanaluk K, Suwancharoen D, Silpasakorn S, Chierakul W, Day N. 2010. Strategies for diagnosis and treatment of suspected leptospirosis: a cost-benefit analysis. PLoS Negl Trop Dis 4:e610. https://doi10.1371/journal.pntd.0000610.

40. Smith S, Liu YH, Carter A, Kennedy BJ, Dermedgoglou A, Poulgrain SS, Paavola MP, Minto TL, Luc M, Hanson J. 2019. Severe leptospirosis in tropical Australia: Optimising intensive care unit management to reduce mortality. PLoS Negl Trop Dis13:e0007929. https://doi10.1371/journal.pntd.0007929

